# Presurgical ablative radiation associates with local control and immune response in pancreatic cancer

**DOI:** 10.1101/2024.11.11.24317120

**Authors:** Peter Q. Leung, Eslam A. Elghonaimy, Ahmed M. Elamir, Megan Wachsmann, Song Zhang, Neha Barrows, Hollis Notgrass, Ethan Johnson, Cheryl Lewis, Rachel von Ebers, Cassandra Hamilton, Grace Josephson, Zhikai Chi, Salwan Al Mutar, Patricio M. Polanco, Nina N. Sanford, Syed M. Ali Kazmi, Matthew R. Porembka, David Hsiehchen, Adam C. Yopp, John Mansour, Muhammad S. Beg, Herbert J. Zeh, Todd A. Aguilera

**Author notes:** **Correspondence:** Todd A. Aguilera M.D. Ph.D. 2201 Inwood Road, NC7.208, Dallas, TX 75390-9187.

## Abstract

**Purpose:** To compare outcomes and molecular characteristics of patients who had surgery after neoadjuvant chemotherapy, with and without ablative radiotherapy (SAbR) for pancreas cancer.

**Experimental Design:** This single-institution, tertiary care academic center cohort study included all patients diagnosed with pancreatic cancer between 2012-2023 treated with neoadjuvant chemotherapy, with or without SAbR. We compared therapeutic responses, performed cardinality matching with distance-optimized pairing, and conducted multivariable stepwise-AIC-optimized Cox modeling to identify differences between groups. We assessed molecular response using RNA sequencing to identify SAbR-induced biologic differences.

**Results:** Among 133 patients receiving chemotherapy and 48 chemotherapy + SAbR, RNA sequencing was available for 29 and 14 patients, respectively. Despite more advanced baseline disease, the SAbR group showed better post-treatment pathology and similar overall survival (HR = 0.97, 95% CI = 0.58–1.60, *P* = .9). Patient matching indicated that SAbR improved locoregional recurrence-free survival (HR = 0.24, 95% CI = 0.07–0.88, *P* = .009). Arterial involvement raised local failure risk with chemotherapy alone (HR = 3.37, 95% CI = 1.74–6.54, *P* < .001), which was significantly reduced with SAbR (HR = 0.28; 95% CI = 0.12–0.68; *P* = .003). Gene set enrichment analysis showed immune activation, with CD8 and NK/NKT cell signatures associated with local control, and Treg signatures associated with worse control.

**Conclusion:** Neoadjuvant SAbR resulted in improved pathological outcomes, enhanced local control, and maintained survival while inducing a distinct immune response. The role of neoadjuvant SAbR should be further evaluated in well powered studies to define clinical benefits.

## INTRODUCTION

Pancreatic ductal adenocarcinoma (PDAC) has a poor prognosis, with surgical resection offering the best chance for 5-year survival—13% overall and 30% for resected cases^1, 2^. Neoadjuvant (NA) therapy helps optimize surgical candidates and may improve local control, as the ESPAC-4 trial highlighted that 50% of failures were locoregional alone, highlighting the critical need for local control^2, 3^. Favorable outcomes from NA therapy for borderline resectable pancreatic cancer (BRPC) were observed in the PREOPANC trials^4–7^. There remains a need to maximize local control, and radiotherapy (RT) may play an essential role in improving local control as systemic therapy improves, especially for unresectable PDAC^8, 9^, the topic of the forthcoming NRG GI011 trial.

The role of NA RT remains unclear. The Alliance A021501 trial showed low resection rates for BRPC patients: 49% in the chemotherapy arm and 35% in the chemotherapy + RT arm^10^. Though not powered for comparison, the RT arm did not meet efficacy criteria. However, the PREOPANC trials and retrospective studies suggest NA RT may improve locoregional control, and patients with positive margins after RT showed similar control to those with negative margins without RT^11, 12^. It is unclear whether RT would benefit the resectable, BRPC, or locally advanced (LAPC) populations most. With local only failure of 50% and local and distant failure in an additional 10% of patients in the ESPAC-4 trial, the importance of locoregional control in resectable disease may be signficant^2^. Furthermore, the RTOG 0848 trial recently reported a survival advantage from adjuvant RT for resectable disease with negative nodes, suggesting RT may improve survival in resectable node negative cases due to local control, while RT can local control for BRPC and LAPC^13^.

Stereotactic ablative radiotherapy (SAbR), also called stereotactic body radiotherapy (SBRT), is designed for tumor ablation through external beam dose escalation. SAbR is becoming more achievable and common in pancreatic cancer, offers shorter treatment duration, reduced toxicity, and potential for improved disease control with dose escalation over conventional chemoradiotherapy^14, 15^. Our multidisciplinary team treats patients with BRPC and LAPC using NA SAbR following 3-6 months of chemotherapy, especially in cases of arterial involvement, aiming to reduce post-surgery locoregional failure. In this NA cohort review, we observed enhanced locoregional control with SAbR, that was pronounced with arterial involvement, alongside increased myeloid and T cell responses, with cytotoxic lymphocytes linked to better local disease control.

## METHODS

### Patient Inclusion

The goal was to evaluate the institutional NA experience that includes chemotherapy alone and chemotherapy + SAbR to gain perspective on the impact of these two approaches despite different patient risk profiles. We identified all patients who received care at UTSW with PDAC diagnoses between October 2012 and February 2023 who received chemotherapy +/- RT before surgical resection. Each chart was verified with manual review.

### Statistical Analysis

Significance thresholds for clinical analyses used a threshold of 0.05. Proportions were tested using Fisher’s exact tests. One-way ANOVA was used to compare means of three or more groups. *P* values for all survival statistics were compared using log-rank tests and Wald tests from Cox modeling.

Recurrence free survival (RFS) and overall survival (OS) were calculated from time of primary tumor resection. Locoregional recurrence was defined by 1) recurrence around the original tumor site (ex. surgical bed) or regional draining lymph nodes on date of imaging detection and 2) medical intervention to control recurrence(s). Distant recurrence has the same definition except recurrence location is at distant sites (ex. liver, lung). Hazard ratios (HR) and their 95% confidence intervals (CI) were calculated from Cox modeling with *P* values from the Wald test. T-tests were assumed with unequal variances (Welch’s). One-to-one cardinality matching^16, 17^ with Mahalanobis-distance-based optimal pairing^18^ with Matchit^19^ R package was used for patient matching: standardized mean difference < 0.015 was used to balance national comprehensive cancer network (NCCN) resectability status, tumor-arterial involvement, tumor-venous involvement, clinical T staging, and time on chemotherapy for each treatment group. Forward selection and backward elimination stepwise Akaike Information Criterion (StepAIC) was used to select the optimized multivariable Cox model. Variables that had no more than 20 missing values were included for optimization. Presurgical characteristics included NCCN resectability status, tumor-arterial involvement, tumor-venous involvement, tumor size, and clinical T staging, all assessed at diagnosis/pre-treatment, as well as sex, chemotherapy regimen, time on chemotherapy, age, and SAbR treatment. Post-surgical pathological characteristics included CAP “Pancreas (Exocrine)” protocol anatomical tumor site, grade, ypT stage, ypN stage, lymphovascular invasion, perineural invasion, treatment effect, and margin status. Adjusted *P* values for selecting GSEA gene sets or differentially expressed genes were adjusted with Benjamini-Hochberg procedure.

### NCCN Anatomical Resectability and Vessel Involvement

Anatomical resectability was defined according to National Comprehensive Cancer Network (NCCN) resectability status guidelines for pancreatic adenocarcinoma (Version 2.2021) defined in real time and documented by the multidisciplinary tumor board and confirmed by radiology report and clinic notes. Tumors were classified as resectable, BRPC, or LAPC based upon NCCN criteria such as the degree of vessel involvement. In cases without multidisciplinary team documentation, the radiology reports and clinic notes of treating oncologists were reviewed for consistency. If there was discrepancy in this review or was a discrepancy from team documentation and chart review, then a board-certified radiation oncologist specialized in pancreatic cancer (Aguilera) confirmed each case based on NCCN criteria. CT scans from pre-treatment, at the initial cancer diagnoses were used for tumor-vessel contact according to these guidelines. Arterial involvement was defined by tumor contact of the superior mesenteric artery, celiac trunk, or common hepatic artery from radiology reports at time of diagnosis. Venous involvement was defined by tumor contact of the superior mesenteric vein or the portal vein upon diagnosis from radiology reports at time of diagnosis.

### Systemic Chemotherapy

The chemotherapy regimen was most commonly FOLFIRINOX or Gemcitabine/nab-paclitaxel depending on patient performance status and medical oncologist discretion. Time on chemotherapy was defined as cycles of chemotherapy multiplied by the standard cycle length of the specific regimen administered as follows: FOLFIRINOX 14 days, Gemcitabine + Abraxane 28 days, Gemcitabine + Capecitabine 28 days, Gemcitabine single agent 28 days, Gemcitabine + Cisplatin 21 days.

### Ablative Radiotherapy

This retrospective evaluation focused on patients simulated supine with arms up in a Stereotactic Frame using a Vacuum bag. Motion management with a breathhold and fiducial markers for tracking was preferred. Multiphasic CT contrast imaging was used to delineate target volumes, often fused with MRI in the simulation position. Image guidance using fiducials or stents accounted for respiratory motion, and motion inclusive Internal Target Volume was determined by evaluating multiple scans during simulation or a 4D scan.

Until 2019 the radiation field included the tumor alone with margin but then added an elective nodal volume most consistent with the triangle volume given emerging evidence that failure beyond the tumor alone is a risk^20–22^. The radiation dose increased from 33 Gy in 5 fractions to 55 Gy in 5 fractions treated two times a week as the team implemented new technologies, gained clinical experience, and enrolled patients on dose escalation studies. This volume did not extend to larger fields like those in RTOG 0848^13, 23^. In cases with critical luminal structures (duodenum, stomach, and bowel), priority was to preserve the constraints, aiming for V33 <1cc and Max 37 Gy with the 3 mm PRV at V35 <1cc and Max 40 Gy. Ideal PTV coverage was 95% of the prescribed dose, with 50-90% being acceptable, and 130% max dose. Patients on clinical trials followed protocol guidelines.

### Surgical resection

Assessment of surgical resectability at our institution was determined by first classifying NCCN resectability criteria then a multidisciplinary discussion with radiology, surgical oncologists, medical oncologists, and radiation oncologists. Decision making was based upon patient tolerance of NA chemotherapy, CA 19-9 response, imaging stability/response, patient comorbidities, and technical considerations for type of surgery and whether it could be performed robotically or open. Radiation was considered for all BRPC and most often pursued if there was arterial abutment. For LAPC there were two types of cases, those with stated plan for exploration prior to radiation, and those who received consolidative SAbR whom the team felt would benefit from surgical exploration. The team aimed to schedule surgery between 4-8 weeks after the end of chemotherapy or radiotherapy.

### CA19-9 laboratory values

The carbohydrate antigen 19-9 (CA 19-9) levels were collected from four key timepoints when available. Pre-chemo/treatment CA19-9 levels are defined as the latest CA19-9 measurement before and within 90 days of the first chemotherapy treatment. Post-chemo CA19-9 is defined as the earliest CA19-9 measurement after the last chemotherapy treatment and no more than 90 days after the last day of chemotherapy treatment. Pre-surgical CA19-9 is defined as the latest CA19-9 measurement after the last chemotherapy treatment and no more than 90 days before surgery. Post-surgery was the first measurement no more than 90 days after surgery. The following four criteria were used to evaluate CA19-9 levels/responses in our multivariable Cox modeling: 1. Log2-normalized proportion changes, 2. return to normal changes (if starting timepoint CA19-9 value was > 38 U/mL and ending timepoint was <= 38 U/mL), 3. whether or not CA19-9 value was normal, and 4. the value of the CA19-9 value at a timepoint itself. Log2-normalized proportion changes and return to normal changes were evaluated for Pre-treatment to post-chemo and presurgical to post-surgical CA19-9 levels.

### Pathological outcomes

Pathological outcomes including post-neoadjuvant/pathological measured (yp) T, ypN, tumor site, margin status, perineural invasion, lymphovascular invasion, histological grade, and treatment effect/response were collected from routine College of American Pathologist “Protocol for the Examination of Specimens from Patients with Carcinoma of the Pancreas” form reports as documented in patient charts at the time of surgical resection.

### RNA extraction and sequencing

Pancreatic tumor surgical specimens were collected from consenting patients. One half of the tumor bed was transected for standard clinical/pathological assessment, and a proportion of the mirror face was stored in RNAlater (Qiagen) and frozen at −80C. Total RNA was then extracted with Qiagen RNeasy Plus Kits for RNA Isolation. RNA concentration and quality were quantified with Nanodrop 2000 UV-VIS Spectrophotometer and Agilent RNA 6000 Pico Kit using Agilent 2100 Bioanalyzer. RNA integrity numbers (RIN) greater than 7.0 were considered acceptable for sequencing. Samples that passed QC were then prepared for sequencing with Illumina Stranded mRNA Prep kit following the manufacturer’s protocol. Briefly, 25-1000ng of total RNA was used as input for poly(A) mRNA enrichment with oligo-dT beads. The enriched mRNA was then fragmented, and first-strand cDNA synthesis was performed using random hexamer primers and reverse transcriptase. Second-strand cDNA synthesis was followed by end repair, A-tailing, and adapter ligation. The libraries were PCR-amplified and purified using AMPure XP beads (Beckman Coulter). The quality and size distribution of the libraries were evaluated using the Agilent 2100 Bioanalyzer. Libraries were quantified using the Qubit dsDNA HS Assay Kit (Thermo Fisher Scientific) and pooled in equimolar amounts. Sequencing was performed on an Illumina NovaSeq 6000 platform targeting 50M reads per sample. Raw sequencing reads were processed using the FastQC (version 0.12.0) tool for quality control. Low-quality reads and adapter sequences were trimmed using Trimmomatic (version 0.32). The clean reads were used with Kallisto (version 0.46.0) to quantify gene expression. TPM abundance was imported into R with tximport (version 1.30.0) package and analyzed with standard DEseq2 (version 1.42.1) pipeline for identification of differentially expressed genes. Gene set enrichment analysis (GSEA) was performed with clusterProfiler (version 4.10.1) package with the Molecular Signatures Database (MSigDB) hallmark(H), ontology (C5), and curated (C2) gene sets.

### Public Datasets for scRNAseq atlas

Singe-cell RNA sequencing (scRNAseq) files from Peng 2019, Steele 2020, Lin 2020, Moncada 2020, and Oh 2023 were used to generate a PDAC scRNAseq atlas^24–28^ **(Supplemental Data Files 1-2)**. Cells were filtered by excluding cells with less than 200 genes, all genes in less than three cells, and genes expressed as being composed of greater than 20% mitochondrial genes. Data were then normalized using the Seurat (version 4.4.0) NormalizeData function with default parameters. Variable genes were detected using the FindVariableFeatures function. Data were scaled and centered using linear regression on the counts and the cell cycle score difference. Principle component analysis was run with the RunPCA function using default parameters. Batch effects were corrected, and samples were integrated by Harmony^29^ using the SeuratWrappers library with default parameters. Cell clusters were identified via the FindNeighbors and FindClusters functions, with 0.8 resolution and UMAP clustering algorithms. A FindAllMarkers table was created, and clusters were defined by canonical markers obtained from the published papers of these databases. T cell and myeloid cells were subsetted and clustered again to generate higher-resolution clustering for more CIBERSORTx and mapping of select gene sets. Gene set scores were added to this atlas with the Addmodulescore function in Seurat.

### PDAC subtype and tumor microenvironment analysis (TME)

Molecular subtyping of PDAC tissues was done using gene signatures derived from public Bailey 2016, Moffit 2015, Collisson & Sadanandam 2011, and George & Kudryashova 2024^30–32^. The Bailey signature was generated using the top 50 DEGs with positive logFC for each subtype vs all others, filtered by the lowest adjusted P values from **Supplemental Data Files 3-6**. Moffit signature was generated from **Supplementary Data File 7** for all genes assigned to Basal-like or Classical subtypes. The Collisson & Sadanandam 62 assigner genes from **Supplementary Data File 8** were used for classical, exocrine-like, and qm-PDA subtypes. ssGSEA was then used to calculate these subtype scores from these three publications, and the samples were assigned to the subtype with the highest score. For the George & Kudryashova TME subtyping, ssGSEA was used to calculate scores for the 25 functional gene sets (representing 4 predefined biologically relevant processes) from **Supplementary Data File 9**. Scores were scaled and z-score normalized before hierarchical clustering of the samples based on these scores into 4 groups. Cumulative distribution function and delta area plots from consensus clustering using hierarchical clustering and Euclidean distance were used to confirm the appropriateness of pre-defining 4 TME subtypes as in George & Kudryashova 2024. Finally, Fisher’s exact test was then used to compare proportions of all these PDAC/TME subtypes in the NA chemotherapy + SAbR vs NA chemotherapy treated samples.

### CIBERSORTx

CIBERSORTx is a computational framework to infer cell type abundance and cell specific gene expression from bulk RNA profiles of intact tissue without the physical isolation of cells^33^. Merged public scRNAseq PDAC datasets were used to create gene signatures and cell proportion estimations through the online modules by the creators of the algorithm. For T-cell subset proportion estimations, all 12,593 high-resolution T-cell subset cells from scRNAseq atlas were used to generate a signature matrix with CIBERSORTx online modules with quantile normalization disabled. The generated custom matrix was used to impute cell fractions without the batch correction, quantile normalization, and absolute mode options. The same was done for the myeloid subset proportion estimations, but a subset of 16,500 representative myeloid cells was used to generate the signature matrix for subsequent imputation of cell fractions for bulk-sequenced samples. Welch’s T tests were used to compare the average proportions of each subset for NA chemotherapy + SAbR vs NA chemotherapy treated samples.

### Scissor

The <u>S</u>ingle-<u>C</u>ell <u>I</u>dentification of <u>S</u>ubpopulations with bulk <u>S</u>ample Phen<u>O</u>type Co<u>R</u>relation (Scissor) algorithm uses bulk-RNA sequenced samples and corresponding/linked patient outcomes to predict which cells on a scRNAseq reference dataset are most associated with the outcomes^34^. High resolution T cell and myeloid subsets from the aforementioned PDAC scRNAseq atlas were used to identify Scissor^34^ cells associated with locoregional recurrence free survival (LR-RFS). An alpha of 0.05 was used without percentage cutoff restriction. The “Cox” response type for regression family was used for identifying Scissor cells. Seurat “FindMarkers” function was used to find differentially expressed genes between cells associated with improved vs worsened local control. Genes with *P* values by Wilcoxon rank sum test < 0.001 and log_2_FC >1.5 were used to create the gene Scissor-identified, local-control-improved gene signature for LR-RFS analysis. ssGSEA assigned this gene signature score for each bulk-RNAseq sample, and corresponding patient LR-RFS was compared for highest-scoring and lowest scoring halves.

### IRB statement

All procedures were conducted under IRB-approved protocol STU 072018-037. Patient consents for tissue collection were performed under IRB-approved protocol STU 102010-098.

### Data availability

In addition to data provided in the supplementary data files, RNA sequencing data is available in the public repository GSE277386 (https://www.ncbi.nlm.nih.gov/geo/query/acc.cgi?acc=GSE277386). Additional deidentified clinical information is available at discretion of the corresponding author.

## RESULTS

### Patient and treatment characteristics

Patients were selected by the multidisciplinary team for resection based on chemotherapy response, tumor markers, and vascular involvement. Those with BRPC and LAPC that had arterial involvement, were considered a high-risk for positive margins and often recommended for SAbR if technically feasible. Fresh tissue from the resection bed was collected for RNAseq from consenting patients **(Figure 1A)**. A total of 133 patients received NA chemotherapy and 48 received NA chemotherapy + SAbR. Tissue for bulk RNAseq analysis was collected from 29 and 14 patients, respectively **(Figure 1B)**. When observing pretreatment features, NA chemotherapy + SAbR had significantly more arterial involvement **(Figure 1C)**, more advanced NCCN resectability status **(Figure 1D)**, longer time on chemotherapy **(Figure 1E)**, and longer time from diagnosis to surgery **(Figure 1F)**. Given that CA19-9 normalization following neoadjuvant therapy has been shown to strongly associated with improved survival outcomes^35^, we ran StepAIC-optimized multivariable Cox modeling on our dataset and observed that normal presurgical CA 19-9 values measured after completion of chemotherapy predicted overall **(Supplemental Figure 1A)** and recurrence free survival **(Supplemental Figure 1B and 1C)** outcomes. All other collected presurgical characteristics are available in **Supplemental Table 1**. The presurgical characteristics of patients in the RNAseq group are shown in **Supplemental Tables 2 and 3**, which are representative of the whole cohort.

**Figure 1:**
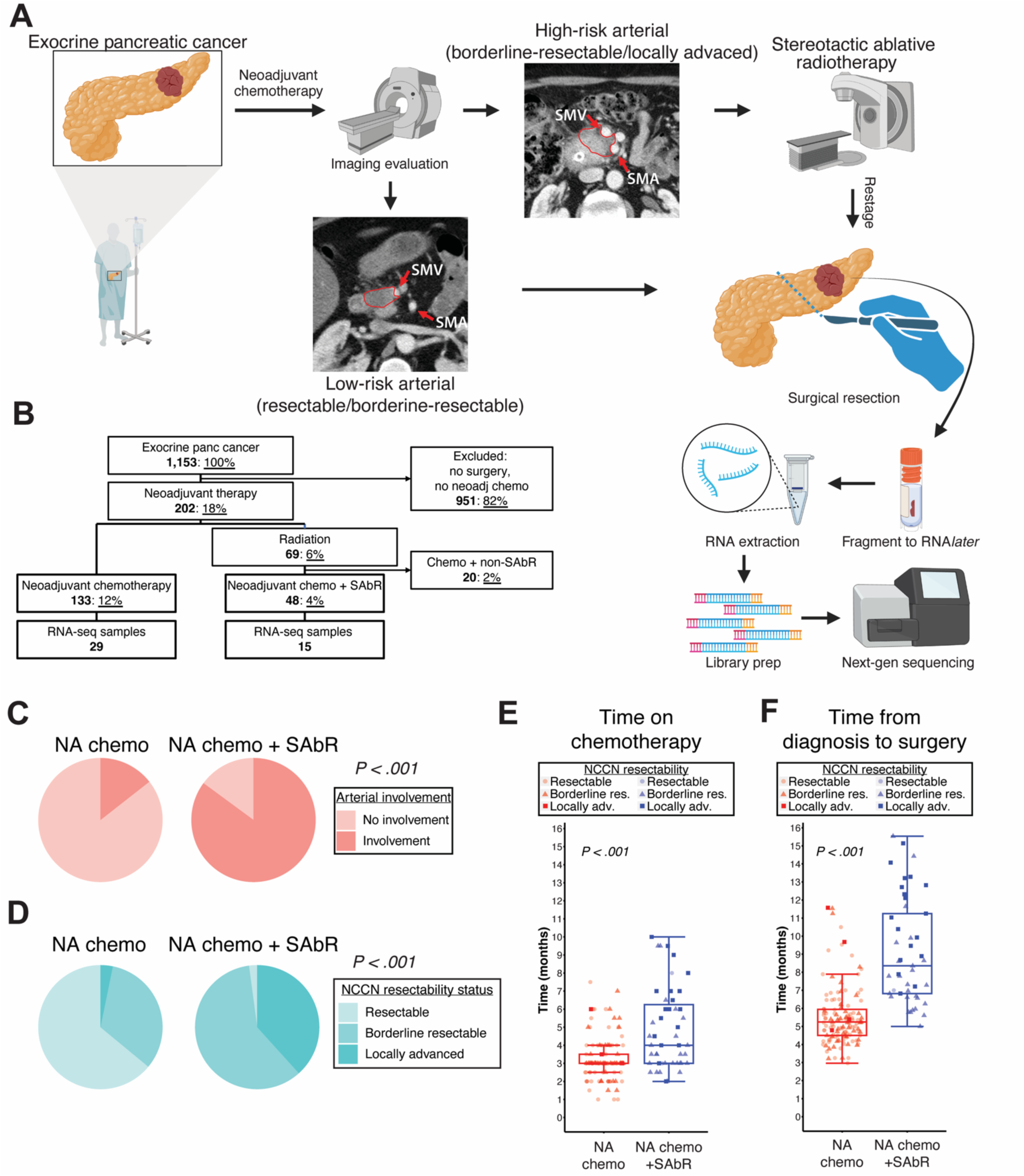
Management of neoadjuvant therapy for PDAC and group characteristics. **(A)** Overview of treatment strategy and tissue collection for this study. **(B)** Patient inclusion/exclusion criteria. **(C-D)** Pie chart proportion representations by neoadjuvant treatment regimens of **(C)** superior mesenteric, common hepatic, or celiac artery involvement at diagnosis by the tumor and of **(D)** resectability based on NCCN criteria. **(E)** Chemotherapy duration, calculated by cycles and standard regimen length. **(F)** Time from confirmed pathological diagnosis to surgical resection. Proportions compared using Fisher’s Exact Test; Welch’s T test used for chemotherapy duration and time to surgery comparisons.

### SAbR results in favorable pathologic outcomes that correlate with survival

To evaluate overall survival outcomes, we first performed patient matching analysis using cardinality matching^16, 17^ with distance-optimized pairing^18^, a computationally robust method to match patients as opposed to propensity score matching, given imbalances of presurgical characteristics **(Supplemental Table 1)**. To reach a high level of balance, 25 patients treated with SAbR and 25 with chemotherapy alone were matched **(Supplemental Table 4)**. For matched patients, the 2-year overall survival of the SAbR-treated group was like those treated with NA chemotherapy alone (55% vs 47%), and univariable Cox analysis of SAbR treatment revealed no significant difference in overall survival (*P* = 0.56) **(Supplemental Figure 2A)**.

Next, we performed multivariable stepAIC-optimized Cox modeling on the whole cohort to determine covariates associated with overall survival. StepAIC optimization selected tumor-venous involvement at diagnosis and pathological T stage as important and significant predictors in the multivariable Cox model for overall survival **(Supplemental Figure 3)**. Of note the receipt of SAbR was not selected nor was it associated with improved survival **(Supplemental Figure 4, 5A)**, but we knew selection of patients for SAbR was biased toward more advanced clinical disease (**Supplemental Table 1**).

Given the multivariable analysis we sought to investigate differences between the NA chemotherapy and NA chemotherapy + SAbR for these and other variables on the whole cohort. At baseline diagnosis, NA SAbR treated patients had a more advanced clinical stage given the high proportion of patients with arterial involvement **(Figure 2A left)**. However, after surgery, the pathologic T stage was in the NA SAbR group was significantly lower than in the NA chemotherapy treated patients (*P* = .002) shown in the alluvial plot and pie charts **(Figure 2A right, B)**. Additionally, pathological ypN stage (*P* = .015), treatment effect (*P* < .001), and perineural invasion (*P* = .019) were significantly improved with SAbR treatment **(Figure 2B-E, top; Supplemental Table 5)**. All of these were correlated with improved overall survival, except perineural invasion **(Figure 2B-E, bottom; Supplemental Table 4)**. In short, these data suggest that improved treatment effect, ypT, and ypN may impact overall survival for patients treated with neoadjuvant therapy.

**Figure 2:**
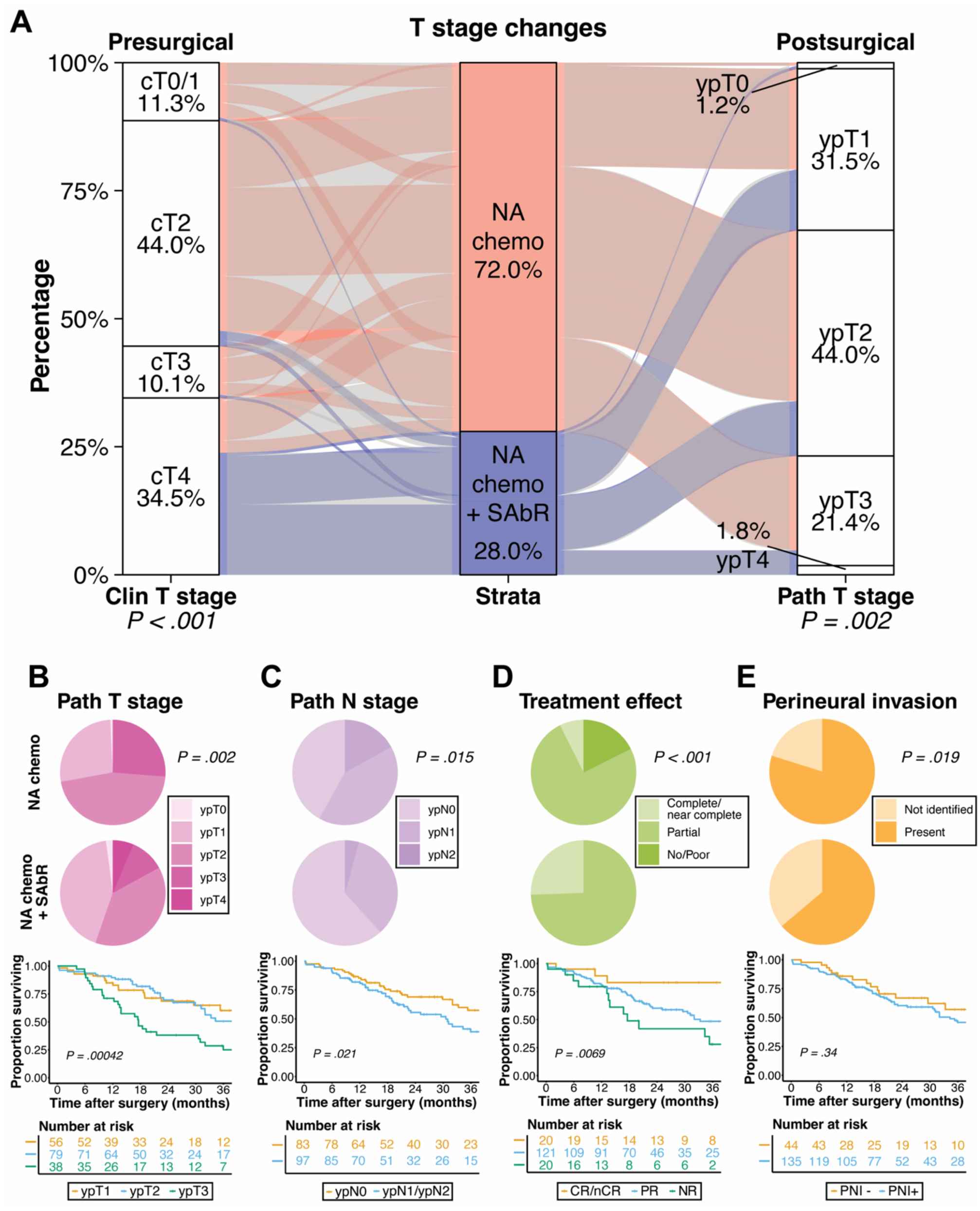
SAbR improves pathological outcomes that correlate with survival. **(A)** Alluvial plot of pre-treatment clinical T stage and post-treatment pathological T stage (ypT) stratified by neoadjuvant treatment regimen. Fisher’s exact test was used to compare proportions within clinical T stage. Fisher’s Exact test was also used for ypT stage. **(B)** Post-treatment ypT-stage proportions by treatment regimen and Kaplan-Meier survival curves (excluding ypT0 and ypT4 due to low patient numbers). **(C)** Post-treatment nodal ypN proportion differences and survival curves based on nodal status for the entire cohort. **(D**) Pathological treatment effect and survival curves as in (**B-C**). **(E)** Perineural invasion proportions and survival curves. Proportion compared using Fisher’s exact test, survival by log-rank test.

Molecular subtyping and analyzing the tumor microenvironment with RNA sequencing could give important insights that may have clinical relevance in PDAC. We first confirmed that patients whose tumors were sequenced were representative of the larger cohort for post-surgical pathological characteristics **(Supplemental Tables 6 and 7)**. We then used Moffitt^31^, Collison & Sadanandam^32^, and Bailey^30^ gene signatures given biological insights gleaned, though these subtypes currently don’t impact clinical management. To explore molecular subtype association with treatment received, we classified patients based on the signatures but found no significant differences between those treated with or without SAbR **(Supplemental Figure 6A-C)**. We next performed transcriptomic subtyping using a recent study’s four PDAC TME phenotypes, ranging from immune enriched to immune depleated^36^. We validated these subtypes using hierarchical clustering into four groups **(Supplemental Figure 6D)**, but did not observe a radiation-induced increase in fibrotic TME subtypes as observed after chemoradiation. This may be because of differences between SAbR and long course chemoradiation. There was no significant subtype proportion changes were observed for different pathologic outcomes either. Because molecular subtypes exist on a continuum, there may be limited ability to detect differences due to the small sample size. Overall, transcriptomic subtypes remained consistent proportion within treatment groups, suggesting further analysis of specific gene pathways or immune response is needed.

### Pathologic features and Myc targets are associated with risk of distant failure

We next aimed to determine how SAbR might impact distant metastasis RFS. First, we used the same subset of 25 NA chemotherapy + SAbR and 25 NA chemotherapy alone treated patients **(Supplemental Table 4)** using cardinality matching^16, 17^ and distance-optimized pairing^18^ to investigate the association of SAbR treatment with distant RFS. The two-year distant RFS was more than double in the SAbR treated matched patients (68% vs 29%), but univariable Cox analysis of SAbR treatment revealed no significant difference in distant RFS (HR = 0.56, 95% CI = 0.26–1.2, *P* = 0.15) **(Supplemental Figure 2B)**.

Given these matched patient results and that the overall cohort of patients treated with NA chemotherapy + SAbR had worse presurgical characteristics (**Figure 1C-F**) yet similar distant failure outcomes **(Supplemental Figure 5B, 7)** compared to those treated with NA chemotherapy, we performed stepAIC-optimization of a multivariable Cox proportional hazards model on the whole cohort to confirm the importance of these variables for predicting distant metastasis RFS. Post-surgical N stage, treatment effect, and lymphovascular invasion were selected as contributing predictors by this method, but not post-surgical T stage **(Figure 3A)**.

**Figure 3:**
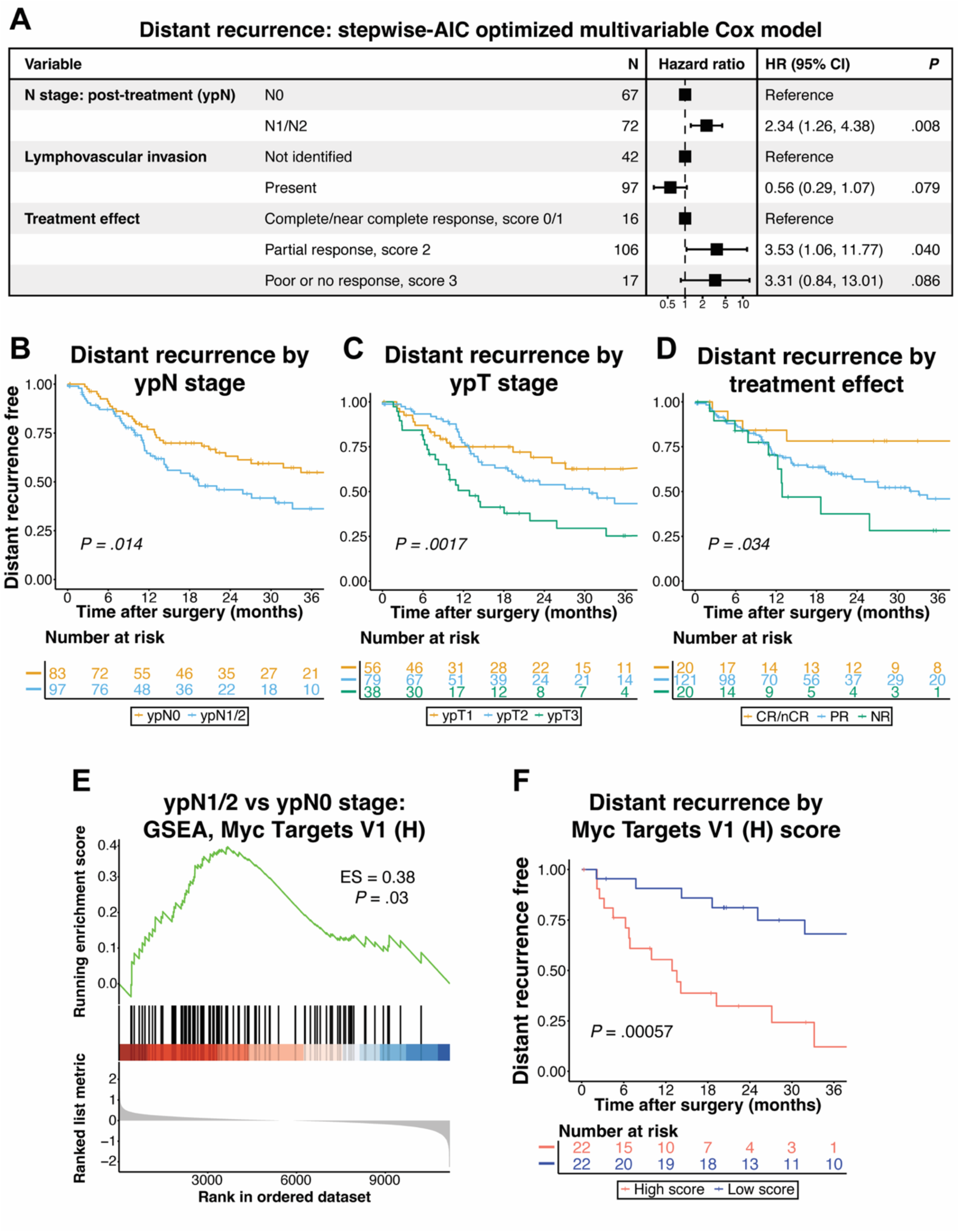
Pathologic features and Myc targets are associated with risk of distant failure. **(A)** StepAIC-optimized Cox model for predicting distant failure. Model considered age at diagnosis, neoadjuvant treatment, tumor site, grade, ypT, ypN, lymphovascular invasion, perineural invasion, treatment effect, clinical T stage, margin status, NCCN resectability status, arterial and venous involvement at diagnosis. Reported *P* values from Wald test. **(B-D)** Distant recurrence free survival Kaplan-Meier curves of all patients that received NA therapy and were resected based on post-treatment pathological **(B)** ypN stage, **(C)** ypT stage, and **(D)** pathologic CAP protocol treatment effect. Reported *P* values from log-rank test. **(E)** GSEA enrichment plot of Hallmark gene set “Myc Targets V1” with Benjamini-Hochberg-adjusted *P* values. **(F)** Kaplan-Meier curve comparing distant metastasis-free survival between high and low “Myc Targets V1” gene set scores via ssGSEA. *P* value calculated from log-rank test.

Given the improved pathological outcomes associated with SAbR, the results of the multivariable analysis, and the association of these post-surgical variables with improved overall survival, we hypothesized similar associations of these factors with distant RFS. Consistent with this hypothesis, we observed an association of improved distant RFS with improved ypN stage (*P* = .014), ypT stage (*P* = .001), and treatment effect (*P* = .034) **(Figure 3B-D).** Univariable Cox models for these factors are summarized in **Supplemental Figure 7**. In summary, distant failure is lower in patients with several improved pathological features, which we showed in **Figure 1** are improved by SAbR despite more advanced clinical stage.

Identifying potential biomarkers for distant failure could be useful in clinical decision making. Therefore, we investigated transcriptomic features that have been associated with distant failure such as transcriptional targets of the *MYC* oncogene^37^. We hypothesized that patients with nodal metastases shown to have greater risk of distant metastases may reveal a transcriptomic link in the primary tumor tissue. Analysis of bulk RNAseq revealed the hallmark gene set “Myc Targets V1” to be significantly upregulated in patients with positive nodal status at surgical resection **(Figure 3E, Supplemental Data File 10)**. We then divided all bulk RNAseq samples into high and low signature score based on this gene set and found patients with a lower score had significantly lower rates of distant metastasis recurrence after surgery compared to those that had a higher “Myc Targets V1” score (HR = 0.30, 95% CI = 0.09– 0.56, log-rank *P* = .00057) **(Figure 3F)**. These results confirm *MYC* signaling in primary resection samples as a potential prognostic indicator of increased likelihood of distant metastasis after surgical resection.

### NA SAbR results in greater local control in PDAC

We hypothesized SAbR could impact locoregional recurrence free survival (RFS) with direct effects on tumor cells and an antitumor immune response. We used the same subset of 25 patients treated with NA chemotherapy + SAbR and 25 NA chemotherapy alone **(Supplemental Table 4)** using cardinality matching^16, 17^ and distance-optimized pairing^18^ to investigate the association of SAbR treatment with local control. The 2-year locoregional RFS of the SAbR-treated group was more than double that of NA chemotherapy alone (78% vs 35%), and Cox model analysis revealed improved locoregional RFS with SAbR treatment (HR = 0.24, 95% CI = 0.07–0.88, *P* = .009) (**Figure 4A**).

**Figure 4:**
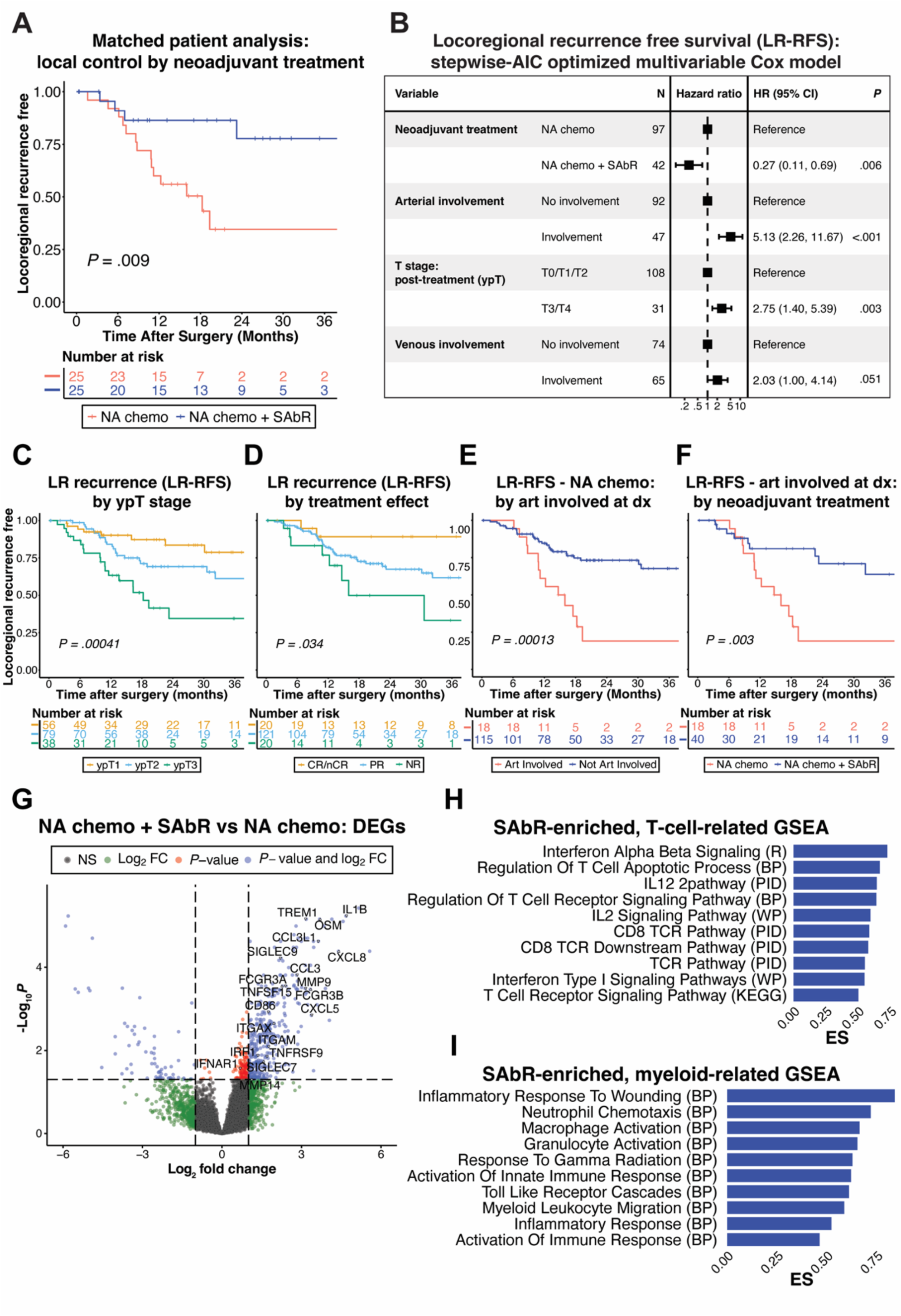
SAbR results in greater local control and immune cell pathway enrichment. **(A)** Matched patient analysis comparing locoregional recurrence free survival (RFS) stratified by neoadjuvant SAbR treatment. **(B)** StepAIC-optimized Cox model for predicting locoregional recurrence considering the same variables as in Figure 3A. *P* values from Wald test. Kaplan-Meier locoregional recurrence free survival curves (LR-RFS) for all patients who received NA therapy stratified by **(C)** ypT and **(D)** treatment effect. **(E)** Kaplan-Meier LR-RFS curves for NA chemotherapy treated patients stratified by tumor arterial involvement at diagnosis. **(F)** Kaplan-Meier LR-RFS curves for patients with arterial involvement by their tumors at diagnosis, stratified by neoadjuvant treatment. All Kaplan-Meier curve *P* values from log-rank tests. **(G)** Differentially expressed genes comparing NA chemotherapy + SAbR treated bulk RNAseq samples to those without SAbR. Select immune-response related genes are highlighted. **(H-I)** Select GSEA gene sets from Reactome (R), Biological Process (BP) subset of C5 Gene Ontology, Pathway Interaction Database (PID) subset of C2 Canonical Pathways, WikiPathways (WP) subset of C2CP, and Kyoto Encyclopedia of Genes and Genomes (KEGG) subset of C2CP databases.

Given that pathological characteristics were correlated overall survival and distant metastasis RFS, we hypothesized there would be similar results with locoregional control. To identify variables that predict locoregional RFS, we performed multivariable stepAIC-optimized Cox modeling on the whole cohort. StepAIC optimization of the Cox regression model confirmed that SAbR treatment, ypT stage, and arterial involvement at diagnosis were significant for predicting improved locoregional recurrence free survival **(Figure 4B)**. Therefore, if SAbR treatment can improve local control, ypT stage and arterial margin there would be important value, especially in those patients that have radiographic arterial involvement at diagnosis.

To explore the differences for patients who did and did not receive SAbR, we observed that the post-surgical T stage and treatment effect correlated with delayed locoregional recurrence (log-rank *P* = 0.00041 and 0.034, respectively) **(Figure 4C-D)**. This is in the context of 46% of patients who received SAbR had ypT0-1 compared to only 28% in the patients treated with chemotherapy alone. A total of 84% of patients with ypT1 tumors were free from locoregional recurrence at 2 years compared to only 35% for those with ypT3 tumors (HR = 0.2241, 95% CI = 0.10–0.51, log-rank *P* = .0011). Similarly, 89% of patients with CR/nCR remained free from locoregional recurrence at 2 years compared to 50% of patients with partial or no response (HR = 0.15, 95% CI = 0.03–0.76, log-rank *P* = .012). These data suggest that SAbR may influence TNM staging factors that determine prognosis, potentially impacting outcomes in cases where ypTNM staging was not established in patients who received treatment.

To evaluate locoregional control, we stratified all patients by arterial involvement at the time of diagnosis. We hypothesized that those that did not receive SAbR would have greater locoregional failure. At 24 months, only 15% of arterial involved NA chemotherapy-treated patients remained free of locoregional recurrence compared to 71% of those patients without arterial involvement **(Figure 4E)** (HR = 3.37, 95% CI = 1.74–6.54, log-rank *P* = .00013). This was not due to differential presurgical characteristics because the groups were balanced except for the features defined by arterial involvement, the NCCN resectability status and clinical T stage **(Supplemental Table 8)**. We next evaluated the patients that received NA chemotherapy + SAbR and observed that at 24 months over 70% of patients with arterial involvement had locoregional control compared to the 15% who only received chemotherapy (HR = 0.28, 95% CI = 0.12–0.68, log-rank *P* = .003) **(Figure 4F)**. The differences between arterial involved NA chemotherapy and NA chemotherapy + SAbR cohorts are unlikely due to differences in presurgical characteristics **(Supplemental Table 9)**. Given the above findings, a summary of the univariable cox proportional hazards models are found in **Supplemental Figure 8.**

### Molecular features differentiating the use of SAbR reveal induced immune signaling

With the observation that SAbR can improve local control of PDAC, we performed differential gene analysis of bulk RNAseq to nominate molecular differences between NA chemotherapy versus NA chemotherapy + SAbR tissue at resection. We observed 597 differentially expressed genes (DEGs), many of which were immune related **(Figure 4G, Supplemental Data File 11)**. To identify biologic features of the DEGs, we performed gene set enrichment analysis (GSEA) and observed that multiple T cell **(Figure 4H)** and myeloid-cell **(Figure 4I)** related gene sets that were significantly upregulated in the NA chemotherapy + SAbR cohort **(Supplemental Data File 12)**. Notably, type I interferon signaling, IL12, and TCR signaling pathways were elevated after SAbR, supporting the notion that RT can enhance adaptive T cell responses^38^. However, T cell response may be countered by myeloid responses including macrophage and granulocyte activation in the setting of multiple immune response gene sets (**Figure 4I**).

Given the evidence of T and myeloid cell changes via GSEA, we sought to explore the specific cell types responsible for these changes. We built a single cell sequencing atlas of over 100,000 cells from 53 samples across 5 PDAC public datasets **(Figure 5A, Supplemental Data Files 1-2)**. We projected the GSEA gene sets from **Figure 4H and 4I** onto this atlas and confirmed the DEGs were largely associated with T and myeloid cells **(Figure 5B)**. We next aimed to determine cell type specific subpopulations from the SAbR-induced gene-expression changes, thus performed CIBERSORTx^33^, a cell proportion deconvolution algorithm, and we used higher resolution T and myeloid cell subsets **(Figures 5C and 5E)** to construct the respective signature matrices. The imputed cell proportion of our bulk RNAseq samples revealed no detectable differences in T cell subset **(Figure 5D)** nor myeloid subset **(Figure 5F)** proportions and broad heterogeneity from patient to patient was observed. In total, these results, along with our observations from TME subtyping analysis **(Supplemental Figure 6)**, indicate cell subset specific phenotypic signaling T and myeloid cells as opposed to cell proportion changes being key to the overall SAbR-induced differences.

**Figure 5:**
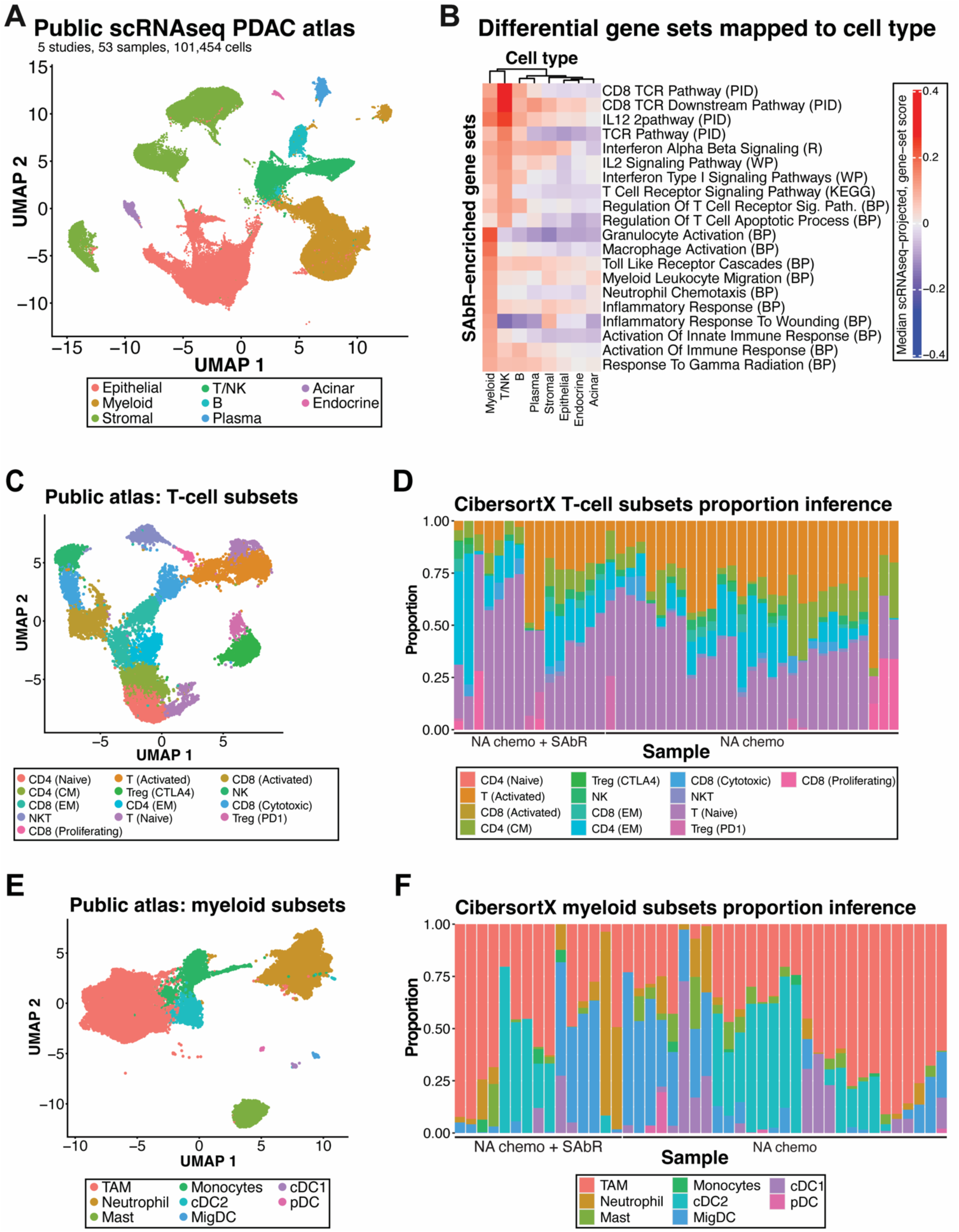
SAbR induced immune signaling is observed in T cells and myeloid cells without changes in cell proportions. **(A)** Single cell RNA sequencing UMAP from public PDAC datasets. **(B)** Median gene set scores for each cluster in scRNAseq atlas. Gene sets are the same SAbR enriched gene sets in Figure 4H-I. **(C)** High-resolution scRNAseq T cell subsets from public PDAC datasets. **(D)** CIBERSORTx relative cell proportion inference of bulk RNAseq, neoadjuvant-treated PDAC samples based on T cell subsets and reference gene signatures built from **C**. **(E)** High-resolution scRNAseq myeloid subsets from public PDAC datasets. **(F)** CIBERSORTx relative cell proportion inference of bulk RNAseq, neoadjuvant-treated PDAC samples based on myeloid subsets and reference gene signatures built from Figure 5E.

### Cytotoxic lymphocytes are associated with improved local control

Given the T and myeloid cell changes observed, we hypothesize that CD8 T cell activation is responsible for local control benefits of SAbR. Therefore, we applied the <u>S</u>ingle-<u>C</u>ell <u>I</u>dentification of <u>S</u>ubpopulations with bulk <u>S</u>ample Phen<u>O</u>type Co<u>R</u>relation (Scissor) algorithm^34^ to determine which T and myeloid cells in the scRNAseq reference atlas correlate to local control outcomes in our patients **(Figure 6A)**. The Scissor algorithm revealed cytotoxic CD8 T, NK, and NKT cells as most associated with improved local control. Meanwhile, Tregs were associated with local failure **(Figure 6B, Supplemental Figure 9)**. There were no cells in the myeloid subsets that were significantly associated with local control. To confirm the importance of cytotoxic lymphocytes for local control, we found 308 differentially expressed genes in the Scissor analysis between improved vs worsened local control **(Figure 6C, Supplemental Data File 13)**. Many genes important for cytotoxic anti-tumor response were observed as top hits, such as *GZMB*, *PRF1*, and *GNLY* **(Figure 6C)**. Lastly, we created a signature of these top DEGs, applied the signature to our bulk-sequenced samples, and observed a locoregional recurrence free survival advantage in patients with a high signature score (HR = 0.2443, 95% CI = 0.06026 - 0.9903, log-rank *P* = 0.035) (**Figure 6D**). In summary, Scissor revealed the importance of cytotoxic lymphocytes in promoting and Treg cells in acting against local control, suggesting the importance of these cell types to SAbR-induced improvements in local control in PDAC.

**Figure 6:**
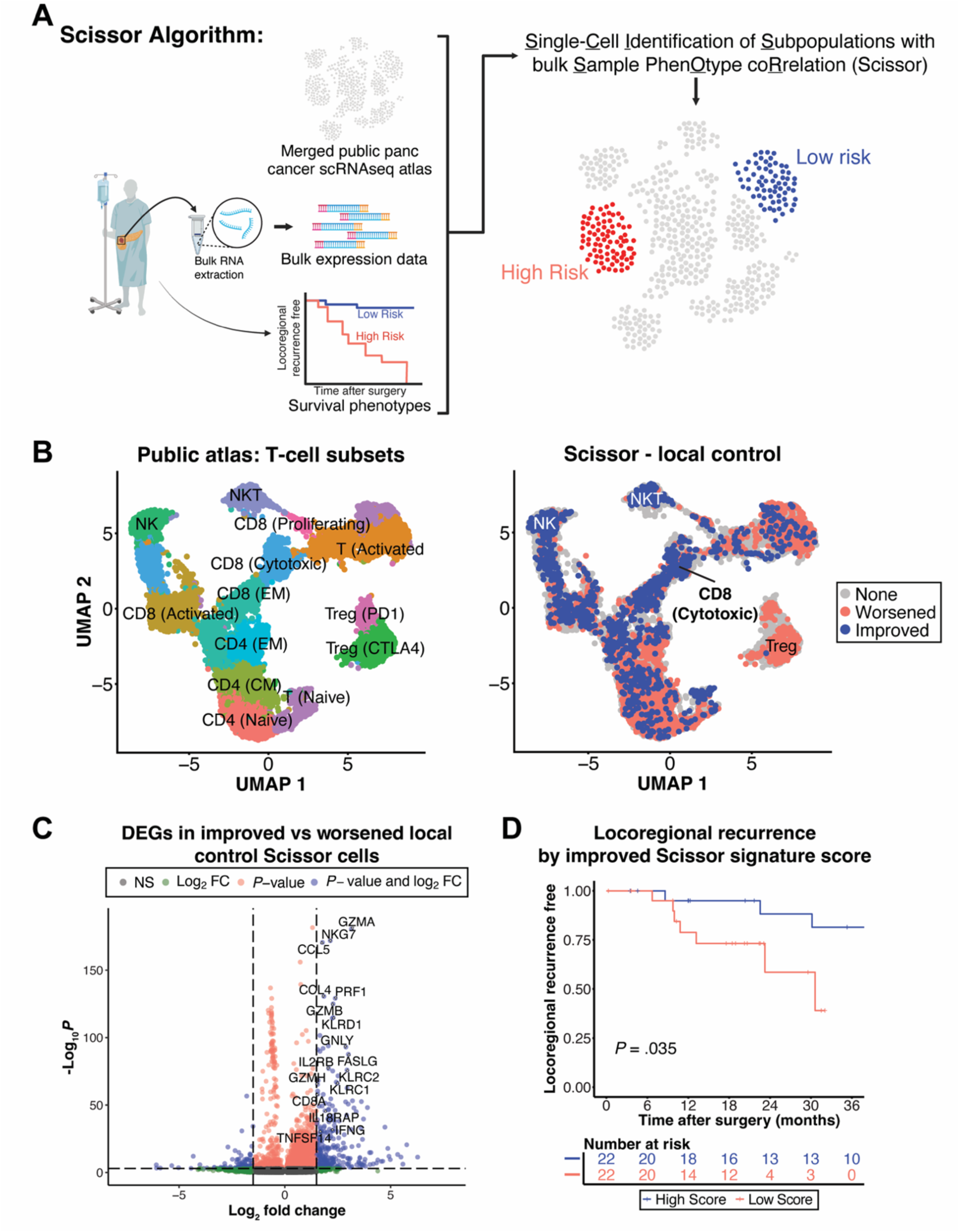
Cytotoxic lymphocytes associate with improved local control, while Tregs associate with worsened local control. **(A)** Overview of the Single-Cell Identification of Subpopulation with bulk Sample Phenotype Correlation (Scissor) algorithm. Inputs of 1. bulk RNAseq samples with 2. corresponding survival phenotypes and a 3. scRNAseq reference atlas into the Scissor algorithm can predict which scRNAseq cells are most correlated and anti-correlated to outcomes. **(B)** High-resolution T cell subsets from public scRNAseq atlas with Scissor-identified cells linked to improved (blue) and worsened (red) local control. NKT, NK, and cytotoxic CD8 T cells correlated the most with improved local control; Tregs with worsened control. **(C)** Differentially expressed genes (DEGs) between Scissor-identified cells for improved vs worsened local control, with cytotoxicity-related DEGs highlighted. **(D)** Kaplan-Meier curve of LR-RFS for patients stratified by gene-expression signature scores from DEGs linked to Scissor-identified cells associated improved local control. DEGs with Log2FC >1.5 and *P* < .001 were used. Kaplan-Meier *P* value from log-rank test.

## DISCUSSION

While evidence suggests neoadjuvant (NA) chemotherapy improves outcomes for patients with pancreatic cancer^4^, the role of NA radiation remains debated^4, 10, 39–42^. Ablative radiation has shown promising survival and disease control in locally advanced disease^8^, and the pending phase III NRG GI 011 trial will evaluate this question. We hypothesize that NA SAbR provides meaningful value and a large well powered study could impact survival given downstaging effects, local control, immune response, and the overall survival benefit observed with adjuvant RT for node negative disease^13^. Our cohort observes these benefits with twice as many resected SAbR-treated patients as the Alliance study, and we observed lower pathologic staging and improved local control compared to patients not treated with RT. Although there are several limitations, this work particularly focused on arterial involvement and the observation of immunologic differences provides important perspective. As suggested by the TAPS consortium study, it will take a large well powered randomized study to definitively show meaningful benefits^42^, which our data supports.

We hypothesize that NA SAbR has three main benefits that are clinically meaningful, first, to improve local control for those with BRPC and LAPC that may extend to beneficial impact beyond improving margins when there is high risk of morbidity from local failure after surgery. Secondly, in all surgical stages, RT may improve survival in the node negative population who are less likely to develop metastases given that survival benefit observed with chemoradiation (CRT) in RTOG 0848^13^. Thirdly, NA SAbR may open opportunities to exploit immunologic vulnerabilities in a disease that has had little impact by immunotherapy. This last point is notable as the SAbR impact on the TME may pay provide a unique opportunity as we better understand this biology given the recently reported randomized EXTEND trial showed systemic immune responses with T cell activation and clonal expansion in patients with PDAC treated with metastasis directed SAbR^43^. These data and others are building a strong case for the systematic evaluation of these questions in patients.

The molecular analysis of resected tissue enables a deeper investigation into heterogeneity and biological response factors. For example, we confirmed that patients with higher MYC target expression have an increased risk of distant failure^37^, which could be an important stratification biomarker for patient selection **(Figure 3E-F)**. Understanding how to assay this preoperatively could aid in surgical decision-making and patient stratification when evaluating the value of aggressive local therapy.

Secondly, in the SAbR group, there was significant T and myeloid cell signaling induced compared to chemotherapy alone. This finding is important given the unclear relationship between radiation fractionation/dose and immune response^44^, especially given PDAC’s limited response to immunotherapies^45^. Thirdly, we did not observe a skewing toward fibrotic TME features as observed in George & Kudryashova et. al. after long course CRT, perhaps from differences between short and long course (**Supplemental Figure 6D**)^36^. Deeper tissue analysis post-SAbR could identify ideal combination therapies. Clinically, consolidating patients stable on chemotherapy with SAbR and immunotherapy could enhance disease control post-chemotherapy, improve outcomes, reduce the toxicity of chemotherapy, and provide immune memory to delay or eliminate recurrence.

This study has several important limitations. First, it is a retrospective review from a high-volume tertiary center, which may affect the generalizability of our findings. Selection bias between factors influencing patients undergoing chemotherapy alone versus the addition of SAbR could also impact treatment response. Additionally, the study focuses on patients that underwent surgery and tissue was not analyzed in patients who did not undergo surgery, thus potentially impacting the interpretation of clinical benefit and tissue assessments. However, recent studies suggest that the majority of patients with BRPC receive NA therapy proceed to resection^46–49^, and resection can be high with initial unresectability^50^. Finally, while we observed improved local control in tumors involving arteries, these findings should provide support for the need to further investigate the value.

In conclusion, we showed tumor arterial involvement at diagnosis is associated with worse locoregional control after NA chemotherapy alone and that the addition of NA SAbR is associated with improved locoregional disease control, in this arterial-involved group and across the entire cohort. With RNA sequencing analysis, we found that SAbR-induced immune-related T and myeloid cell activation changes, with cytotoxic cells correlating with this better disease control. In addition, despite worse pre-operative characteristics, patients treated with NA chemotherapy + SAbR had similar survival outcomes and more favorable pathological outcomes compared to those treated with NA chemotherapy alone. To formally show and establish NA SAbR’s benefits for molecular responses, pathological outcomes, and improved locoregional control, a larger, well-powered, and controlled study of tissue and clinical outcomes in resectable to locally advanced disease is warranted.

## AUTHOR CONTRIBUTIONS

**P.Q. Leung:** Conceptualization, Data curation, Formal Analysis, Investigation, Methodology, Project administration, Software, Validation, Visualization, Writing – original draft, Writing – review & editing. **E.A. Elghonaimy:** Conceptualization, Data curation, Formal Analysis, Investigation, Methodology, Project administration, Resources, Software, Validation, Visualization, Writing – original draft, Writing – review & editing. **A.M. Elamir:** Conceptualization, Data curation, Formal Analysis, Investigation, Methodology, Project administration, Validation, Visualization, Writing – original draft, Writing – review & editing. **M. Wachsmann:** Methodology, Resources, Writing – review & editing. **S. Zhang:** Formal Analysis, Methodology, Validation, Writing – review & editing. **N. Barrows:** Data curation **H. Notgrass:** Resources. **E. Johnson:** Resources, **C. Lewis:** Methodology, Resources. **R. von Ebers:** Data curation **C. Hamilton:** Data curation, Resources. **G. Josephson:** Data curation. **Z. Chi:** Methodology, Resources, Writing – review & editing. **S. Al Mutar:** Methodology, Resources, Writing – review & editing. **P.M. Polanco:** Methodology, Resources, Writing – review & editing. **N. N. Sanford:** Methodology, Resources, Writing – review & editing. **S.M.A. Kazmi:** Methodology, Resources, Writing – review & editing. **M.R. Porembka:** Methodology, Resources, Writing – review & editing. **D. Hsiehchen:** Methodology, Resources, Writing – review & editing. **A.C. Yopp:** Methodology, Resources, Writing – review & editing. **J. Mansour:** Methodology, Resources, Writing – review & editing. **M.S. Beg:** Methodology, Resources, Writing – review & editing. **H.J. Zeh:** Methodology, Resources, Writing – review & editing. **T.A. Aguilera:** Conceptualization, Data curation, Funding acquisition, Methodology, Project administration, Resources, Supervision, Validation, Visualization, Writing – original draft, Writing – review & editing.

## ACKNOWLEDGEMENTS

This research work is supported by the CPRIT Recruitment of First-Time, Tenure-Track Faculty Members RR170051 and the UTSW SCCC Translational Cancer Research Award. We acknowledge the Harold C. Simmons Comprehensive Cancer Center (SCCC) Tissue Management Shared Resource, Clinical Data Exchange Network (ClinDEN) team, UTSW Tumor Registry team. We additionally acknowledge the Dr. Jeon Lee and the SCCC Data Science Shared Resource for training through the Data Science Fellowship. **Figure 1A** and **Figure 6A** were created with Biorender.com and modified with Adobe Illustrator. OpenAI’s ChatGPT large language model (all versions) were used from the date of first release to submission of the manuscript to confirm proper language, improve conciseness, and enhance clarity: authors take full responsibility for the integrity of generated content.

## Transcript Profiling

All RNA sequencing data will be made available in the public repository GSE277386 upon publication of the manuscript, pending acceptance.

## Writing Assistance

No writing assistance was received.

## Data Transparency Statement

Deidentified sequencing characteristics methodology will be made upon reasonable request. All RNA sequencing is available at GEO https://www.ncbi.nlm.nih.gov/geo/query/acc.cgi?acc=GSE277386, will be made available publicly upon publication of the manuscript.

## Notes

**Conflict of interest/ disclosure statement:** All authors have no potential conflicts (financial, professional, or personal) to disclose.

### Competing Interest Statement

The authors have declared no competing interest.

### Author Declarations

IRB of the University of Texas Southwestern gave ethical approval for this work. All procedures were conducted under IRB-approved protocol STU 072018-037. Patient consents for tissue collection were performed under IRB- approved protocol STU 102010-098.

## REFERENCES

1. Siegel RL, Giaquinto AN, Jemal A. Cancer statistics, 2024. CA Cancer J Clin 2024;74:12–49.

2. Neoptolemos JP, Palmer DH, Ghaneh P, et al. Comparison of adjuvant gemcitabine and capecitabine with gemcitabine monotherapy in patients with resected pancreatic cancer (ESPAC-4): a multicentre, open-label, randomised, phase 3 trial. Lancet 2017;389:1011–1024.

3. Strobel O, Neoptolemos J, Jager D, et al. Optimizing the outcomes of pancreatic cancer surgery. Nat Rev Clin Oncol 2019;16:11–26.

4. Versteijne E, Dam JLv, Suker M, et al. Neoadjuvant Chemoradiotherapy Versus Upfront Surgery for Resectable and Borderline Resectable Pancreatic Cancer: Long-Term Results of the Dutch Randomized PREOPANC Trial. Journal of Clinical Oncology 2022;40:1220--1230.

5. Versteijne E, Suker M, Groothuis K, et al. Preoperative Chemoradiotherapy Versus Immediate Surgery for Resectable and Borderline Resectable Pancreatic Cancer: Results of the Dutch Randomized Phase III PREOPANC Trial. J Clin Oncol 2020;38:1763–1773.

6. Janssen QP, van Dam JL, Bonsing BA, et al. Total neoadjuvant FOLFIRINOX versus neoadjuvant gemcitabine-based chemoradiotherapy and adjuvant gemcitabine for resectable and borderline resectable pancreatic cancer (PREOPANC-2 trial): study protocol for a nationwide multicenter randomized controlled trial. BMC Cancer 2021;21:300.

7. Groot Koerkamp B, Janssen QP, Van Dam JL, et al. LBA83 Neoadjuvant chemotherapy with FOLFIRINOX versus neoadjuvant gemcitabine-based chemoradiotherapy for borderline resectable and resectable pancreatic cancer (PREOPANC-2): A multicenter randomized controlled trial. Annals of Oncology 2023;34:S1323.

8. Reyngold M, O’Reilly EM, Varghese AM, et al. Association of Ablative Radiation Therapy With Survival Among Patients With Inoperable Pancreatic Cancer. JAMA Oncol 2021;7:735–738.

9. Parikh PJ, Lee P, Low DA, et al. A Multi-Institutional Phase 2 Trial of Ablative 5-Fraction Stereotactic Magnetic Resonance-Guided On-Table Adaptive Radiation Therapy for Borderline Resectable and Locally Advanced Pancreatic Cancer. Int J Radiat Oncol Biol Phys 2023;117:799–808.

10. Katz MHG, Shi Q, Meyers J, et al. Efficacy of Preoperative mFOLFIRINOX vs mFOLFIRINOX Plus Hypofractionated Radiotherapy for Borderline Resectable Adenocarcinoma of the Pancreas: The A021501 Phase 2 Randomized Clinical Trial. JAMA Oncol 2022;8:1263–1270.

11. Sekigami Y, Michelakos T, Fernandez-Del Castillo C, et al. Intraoperative Radiation Mitigates the Effect of Microscopically Positive Tumor Margins on Survival Among Pancreatic Adenocarcinoma Patients Treated with Neoadjuvant FOLFIRINOX and Chemoradiation. Ann Surg Oncol 2021;28:4592–4601.

12. Zhang E, Wang L, Shaikh T, et al. Neoadjuvant Chemoradiation Impacts the Prognostic Effect of Surgical Margin Status in Pancreatic Adenocarcinoma. Ann Surg Oncol 2022;29:354–363.

13. Abrams RA, Winter KA, Goodman KA, et al. NRG Oncology/RTOG 0848: Results after adjuvant chemotherapy +/- chemoradiation for patients with resected periampullary pancreatic adenocarcinoma (PA). Journal of Clinical Oncology 2024;42:4005–4005.

14. Krishnan S, Chadha AS, Suh Y, et al. Focal Radiation Therapy Dose Escalation Improves Overall Survival in Locally Advanced Pancreatic Cancer Patients Receiving Induction Chemotherapy and Consolidative Chemoradiation. Int J Radiat Oncol Biol Phys 2016;94:755–65.

15. Rudra S, Jiang N, Rosenberg SA, et al. Using adaptive magnetic resonance image-guided radiation therapy for treatment of inoperable pancreatic cancer. Cancer Med 2019;8:2123–2132.

16. Zubizarreta JR, Paredes RD, Rosenbaum PR. Matching for Balance, Pairing for Heterogeneity in an Observational Study of the Effectiveness of for-Profit and Not-for-Profit High Schools in Chile. Annals of Applied Statistics 2014;8:204–231.

17. Niknam BA, Zubizarreta JR. Using Cardinality Matching to Design Balanced and Representative Samples for Observational Studies. JAMA 2022;327:173–174.

18. Hansen BB, Klopfer SO. Optimal full matching and related designs via network flows. Journal of Computational and Graphical Statistics 2006;15:609–627.

19. Ho DE, Imai K, King G, et al. MatchIt: Nonparametric Preprocessing for Parametric Causal Inference. Journal of Statistical Software 2011;42.

20. Kharofa J, Mierzwa M, Olowokure O, et al. Pattern of Marginal Local Failure in a Phase II Trial of Neoadjuvant Chemotherapy and Stereotactic Body Radiation Therapy for Resectable and Borderline Resectable Pancreas Cancer. Am J Clin Oncol 2019;42:247–252.

21. Hill CS, Fu W, Hu C, et al. Location, Location, Location: What Should be Targeted Beyond Gross Disease for Localized Pancreatic Ductal Adenocarcinoma? Proposal of a Standardized Clinical Tumor Volume for Pancreatic Ductal Adenocarcinoma of the Head: The "Triangle Volume". Pract Radiat Oncol 2022;12:215–225.

22. Zhu X, Ju X, Cao Y, et al. Patterns of Local Failure After Stereotactic Body Radiation Therapy and Sequential Chemotherapy as Initial Treatment for Pancreatic Cancer: Implications of Target Volume Design. Int J Radiat Oncol Biol Phys 2019;104:101–110.

23. Goodman KA, Regine WF, Dawson LA, et al. Radiation Therapy Oncology Group consensus panel guidelines for the delineation of the clinical target volume in the postoperative treatment of pancreatic head cancer. Int J Radiat Oncol Biol Phys 2012;83:901–8.

24. Peng J, Sun BF, Chen CY, et al. Single-cell RNA-seq highlights intra-tumoral heterogeneity and malignant progression in pancreatic ductal adenocarcinoma. Cell Res 2019;29:725–738.

25. Steele NG, Carpenter ES, Kemp SB, et al. Multimodal Mapping of the Tumor and Peripheral Blood Immune Landscape in Human Pancreatic Cancer. Nat Cancer 2020;1:1097–1112.

26. Lin W, Noel P, Borazanci EH, et al. Single-cell transcriptome analysis of tumor and stromal compartments of pancreatic ductal adenocarcinoma primary tumors and metastatic lesions. Genome Med 2020;12:80.

27. Moncada R, Barkley D, Wagner F, et al. Integrating microarray-based spatial transcriptomics and single-cell RNA-seq reveals tissue architecture in pancreatic ductal adenocarcinomas. Nat Biotechnol 2020;38:333–342.

28. Oh K, Yoo YJ, Torre-Healy LA, et al. Coordinated single-cell tumor microenvironment dynamics reinforce pancreatic cancer subtype. Nat Commun 2023;14:5226.

29. Korsunsky I, Millard N, Fan J, et al. Fast, sensitive and accurate integration of single-cell data with Harmony. Nat Methods 2019;16:1289–1296.

30. Bailey P, Chang DK, Nones K, et al. Genomic analyses identify molecular subtypes of pancreatic cancer. Nature 2016;531:47–52.

31. Moffitt RA, Marayati R, Flate EL, et al. Virtual microdissection identifies distinct tumor- and stroma-specific subtypes of pancreatic ductal adenocarcinoma. Nat Genet 2015;47:1168–78.

32. Collisson EA, Sadanandam A, Olson P, et al. Subtypes of pancreatic ductal adenocarcinoma and their differing responses to therapy. Nat Med 2011;17:500–3.

33. Newman AM, Steen CB, Liu CL, et al. Determining cell type abundance and expression from bulk tissues with digital cytometry. Nat Biotechnol 2019;37:773–782.

34. Sun D, Guan X, Moran AE, et al. Identifying phenotype-associated subpopulations by integrating bulk and single-cell sequencing data. Nat Biotechnol 2022;40:527–538.

35. Tsai S, George B, Wittmann D, et al. Importance of Normalization of CA19-9 Levels Following Neoadjuvant Therapy in Patients With Localized Pancreatic Cancer. Ann Surg 2020;271:740–747.

36. George B, Kudryashova O, Kravets A, et al. Transcriptomic-Based Microenvironment Classification Reveals Precision Medicine Strategies for Pancreatic Ductal Adenocarcinoma. Gastroenterology 2024;166:859–871 e3.

37. Maddipati R, Norgard RJ, Baslan T, et al. MYC Levels Regulate Metastatic Heterogeneity in Pancreatic Adenocarcinoma. Cancer Discov 2022;12:542–561.

38. McLaughlin M, Patin EC, Pedersen M, et al. Inflammatory microenvironment remodelling by tumour cells after radiotherapy. Nat Rev Cancer 2020;20:203–217.

39. de Geus SW, Eskander MF, Bliss LA, et al. Neoadjuvant therapy versus upfront surgery for resected pancreatic adenocarcinoma: A nationwide propensity score matched analysis. Surgery 2017;161:592–601.

40. Gamboa AC, Lee RM, Maithel SK. The role of radiation for pancreatic adenocarcinoma. Journal of Pancreatology 2020;3:72–80.

41. Lutfi W, Talamonti MS, Kantor O, et al. Neoadjuvant external beam radiation is associated with No benefit in overall survival for early stage pancreatic cancer. Am J Surg 2017;213:521–525.

42. Janssen QP, van Dam JL, Prakash LR, et al. Neoadjuvant Radiotherapy After (m)FOLFIRINOX for Borderline Resectable Pancreatic Adenocarcinoma: A TAPS Consortium Study. J Natl Compr Canc Netw 2022;20:783–791 e1.

43. Ludmir EB, Sherry AD, Fellman BM, et al. Addition of Metastasis-Directed Therapy to Systemic Therapy for Oligometastatic Pancreatic Ductal Adenocarcinoma (EXTEND): A Multicenter, Randomized Phase II Trial. J Clin Oncol 2024:JCO2400081.

44. Demaria S, Guha C, Schoenfeld J, et al. Radiation dose and fraction in immunotherapy: one-size regimen does not fit all settings, so how does one choose? J Immunother Cancer 2021;9.

45. Ullman NA, Burchard PR, Dunne RF, et al. Immunologic Strategies in Pancreatic Cancer: Making Cold Tumors Hot. J Clin Oncol 2022;40:2789–2805.

46. Brown ZJ, Heh V, Labiner HE, et al. Surgical resection rates after neoadjuvant therapy for localized pancreatic ductal adenocarcinoma: meta-analysis. Br J Surg 2022.

47. Masui T, Nagai K, Anazawa T, et al. Impact of neoadjuvant intensity-modulated radiation therapy on borderline resectable pancreatic cancer with arterial abutment; a prospective, open-label, phase II study in a single institution. BMC Cancer 2022;22:119.

48. Barnes CA, Chavez MI, Tsai S, et al. Survival of patients with borderline resectable pancreatic cancer who received neoadjuvant therapy and surgery. Surgery 2019;166:277–285.

49. Javed AA, Wright MJ, Siddique A, et al. Outcome of Patients with Borderline Resectable Pancreatic Cancer in the Contemporary Era of Neoadjuvant Chemotherapy. J Gastrointest Surg 2019;23:112–121.

50. Ferrone CR, Marchegiani G, Hong TS, et al. Radiological and surgical implications of neoadjuvant treatment with FOLFIRINOX for locally advanced and borderline resectable pancreatic cancer. Ann Surg 2015;261:12–7.

