## Supplementary material for "Presurgical ablative radiation associates with local control and immune response in pancreatic cancer": Supplmental figures and tables

**SUPPLEMENTAL FIGURES**

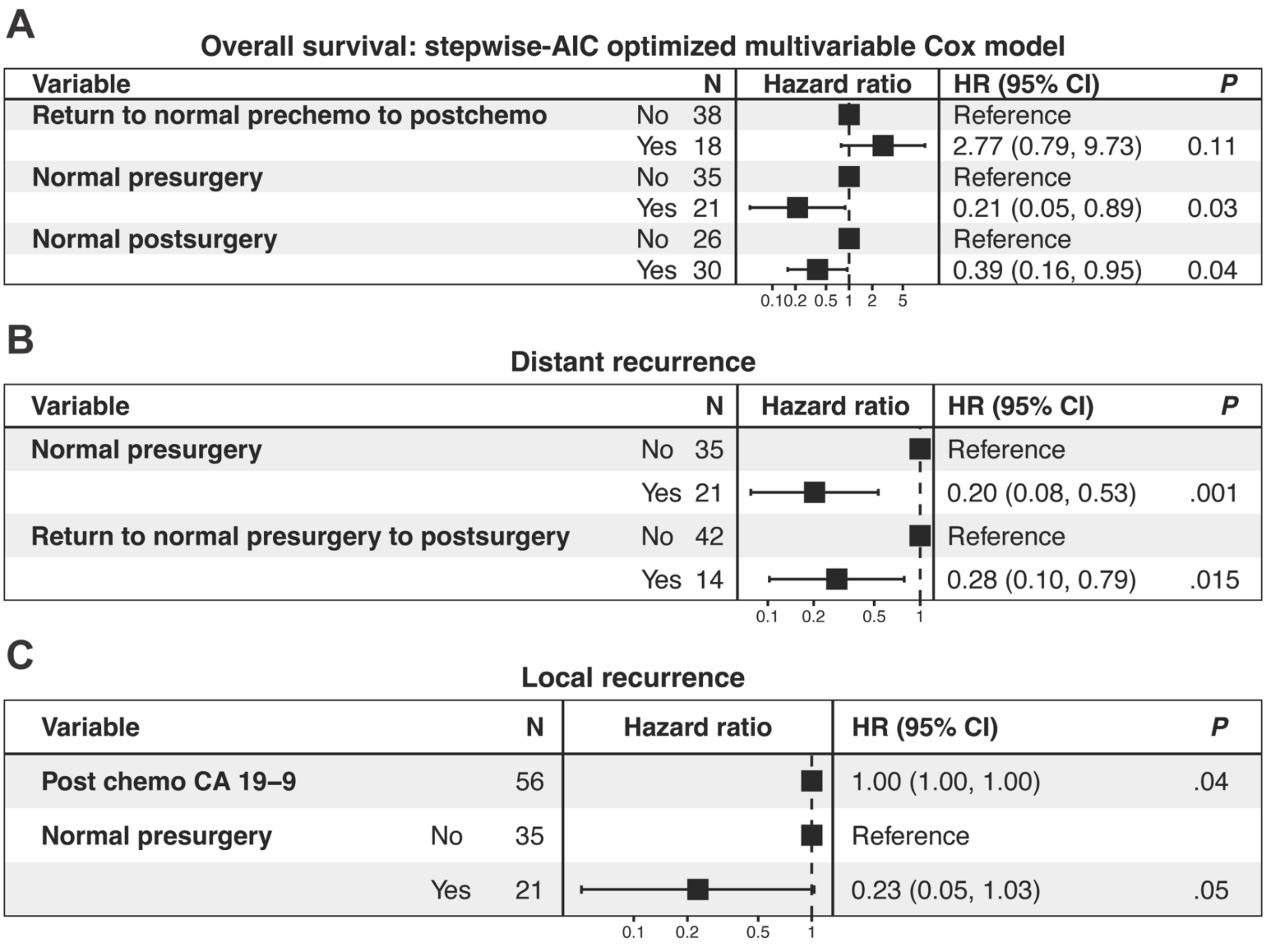


***Supplementary Figure 1: StepAIC-optimized multivariable cox model focusing on CA19-9 characteristics for predicting overall and recurrence-free survival.*** Forward selection and backward elimination Stepwise Akaike information criterion (stepAIC) -optimized Cox proportional hazards models for predicting **(A)** overall **(B)** locoregional recurrence free, and **(C)** distant recurrence free survivals with focus on CA 19-9 values. Model considered pre-chemo CA 19-9 values, post-chemo values, pre-surgery values, post-surgery values, Log2 proportion change (pre-chemo to post-chemo, pre-surgery to post-surgery) as well as binary indicators for returning to normal (pre-chemo to post-chemo, pre-surgery to post-surgery), normal pre-chemo, normal post-chemo, normal pre-surgery, and normal post-surgery. Reported *P* values from Wald test. HR = hazard ratio. CI = confidence interval

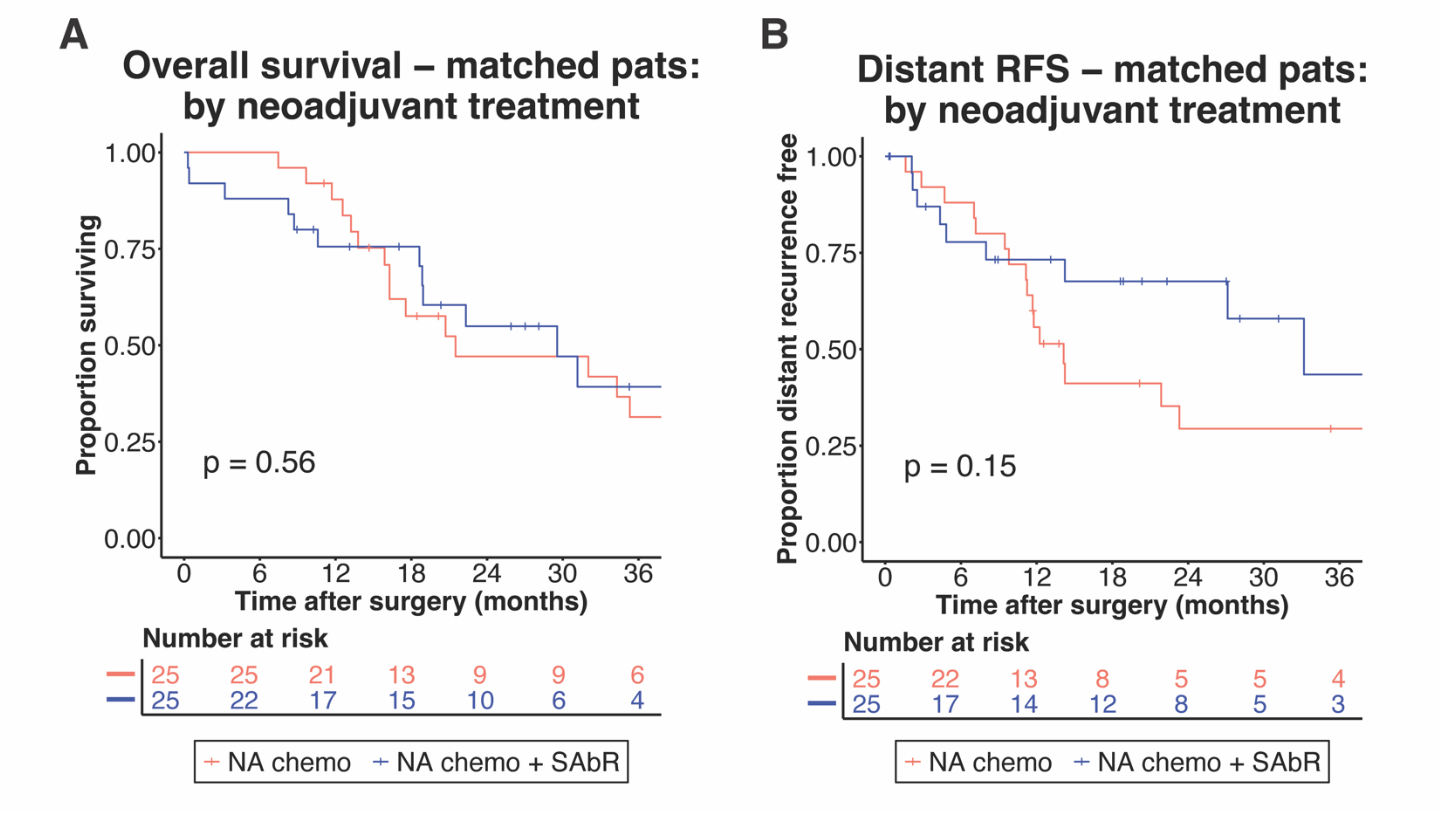


***Supplemental Figure 2: Survival outcomes curves by neoadjuvant treatment regimen for matched patients***.

Kaplan-Meier curves showing **(A)** overall survival, **(B)** distant recurrence free survival of patients treated with NA chemo vs NA chemo + SAbR. *P* values from log-rank test. Patients were matched with cardinality matching with Mahalanobis distance optimized pairing. A standard mean deviation maximum of 0.015 (for covariates of presence tumor-arterial involvement at diagnosis, National comprehensive cancer network (NCCN) resectability status, clinical T stage (AJCC 8th edition), total time on chemo (calculated by total cycles), and presence tumor-venous involvement at diagnosis) was used to achieve maximum patient cohort size and covariate balance.

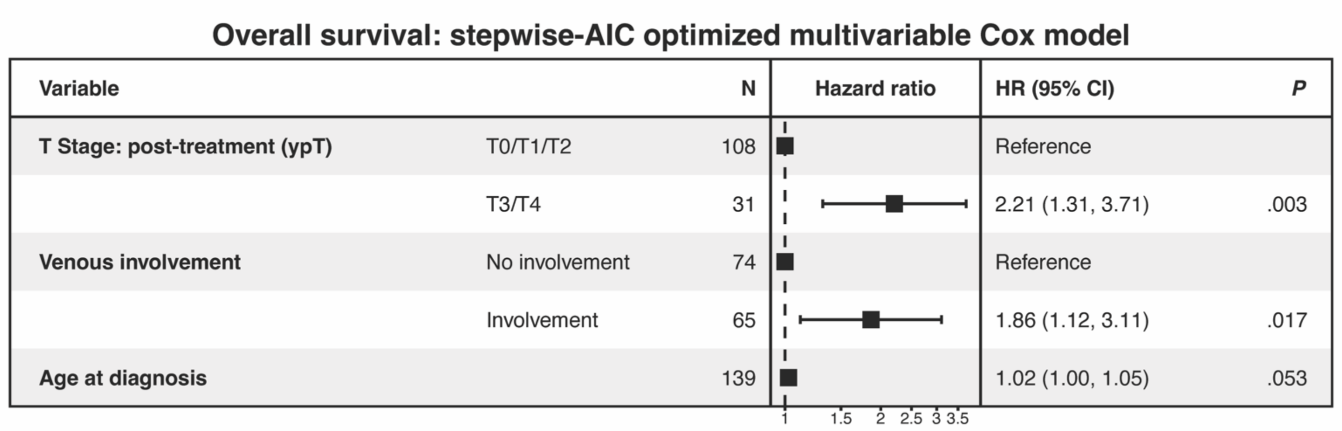


***Supplementary Figure 3: StepAIC-optimized multivariable cox model for predicting overall survival***. Forward selection and backward elimination stepAIC-optimized multivariate Cox model for predicting overall survival. Variables that had no more than 20 missing values were included for optimization. Presurgical characteristics included NCCN resectability status, tumor-arterial involvement, tumor-venous involvement, tumor size, and clinical T staging, all assessed at diagnosis/pre-treatment, as well as sex, chemotherapy regimen, time on chemotherapy, age, and SAbR treatment. Post-surgical pathological characteristics included CAP “Pancreas (Exocrine)” protocol values of anatomical tumor site, grade, ypT stage, ypN stage, lymphovascular invasion, perineural invasion, treatment effect, and margin status. Reported *P* values from Wald test. HR = hazard ratio. CI = confidence interval

### Univariable overall survival Cox models for select variables

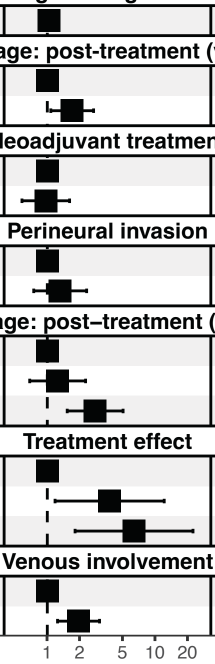

| Variable | N | Hazard ratio | HR (95% CI) | *P* |
|---|---|---|---|---|
| **Age at diagnosis** | | | | |
| Age | 181 | | 1.03 (1.01 – 1.06) | .008 |
| **N stage: post-treatment (ypN)** | | | | |
| No nodal involvement | 83 | | Reference | |
| Nodal involvement | 97 | | 1.70 (1.08 – 2.68) | .022 |
| **Neoadjuvant treatment** | | | | |
| Neoadjuvant chemo | 133 | | Reference | |
| Neoadjuvant chemo + SAbR | 48 | | 0.97 (0.58 – 1.61) | .900 |
| **Perineural invasion** | | | | |
| Not identified | 44 | | Reference | |
| Present | 135 | | 1.32 (0.75 – 2.31) | .340 |
| **T stage: post–treatment (ypT)** | | | | |
| T1: <=2cm greatest dimension | 56 | | Reference | |
| T2: >2cm, <=4 cm greatest dimension | 79 | | 1.25 (0.69 – 2.26) | .468 |
| T3: >4cm greatest dimension | 38 | | 2.77 (1.53 – 5.03) | <.001 |
| **Treatment effect** | | | | |
| Complete/near complete response, score 0/1 | 20 | | Reference | |
| Partial response, score 2 | 121 | | 3.78 (1.18 – 12.11) | .025 |
| Poor or no response, score 3 | 20 | | 6.36 (1.81 – 22.37) | .004 |
| **Venous involvement** | | | | |
| No involvement | 99 | | Reference | |
| Involvement | 82 | | 1.95 (1.24 – 3.05) | .004 |

***Supplementary Figure 4: Univariable Cox model for select variables for predicting overall survival***. Select univariable cox models for variables for predicting overall survival. Variables of interest include neoadjuvant treatment regimen, post-surgical pathological outcomes that were significantly different between NA chemo vs NA chemo + SAbR groups, and those selected with stepAIC in supplementary figure 2. Reported *P* values from Wald test. HR = hazard ratio. CI = confidence interval.

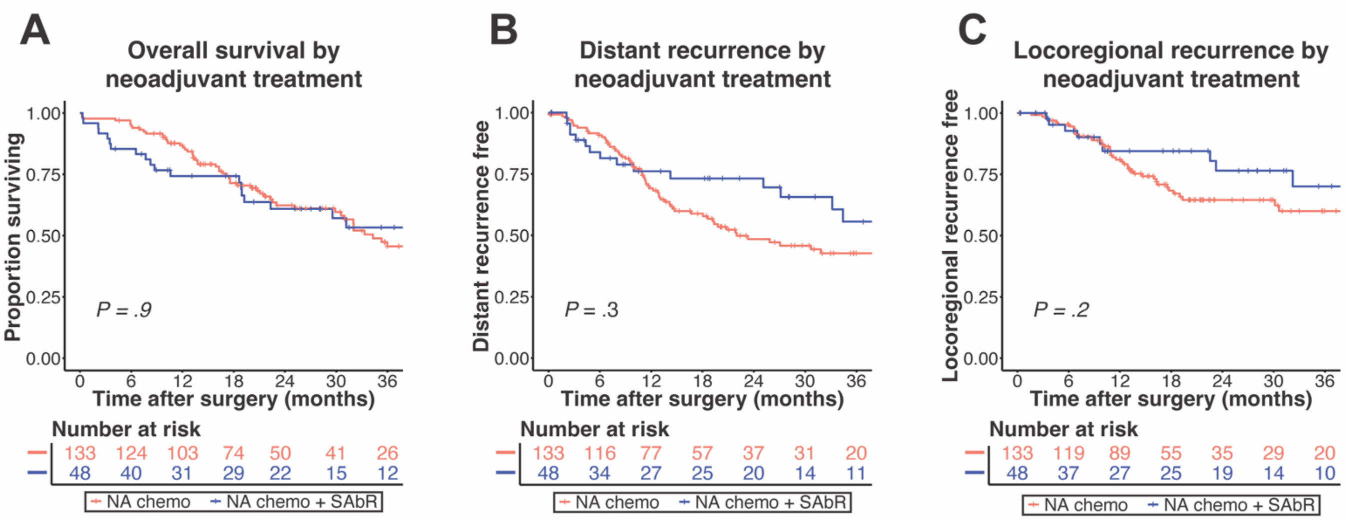


***Supplemental Figure 5: Survival outcomes curves by neoadjuvant treatment regimen for whole cohort***.

Kaplan-Meier curves showing **(A)** overall survival, **(B)** distant recurrence free survival, and **(C)** locoregional recurrence free survival of patients treated with NA chemo vs NA chemo + SAbR. *P* values from log-rank test.

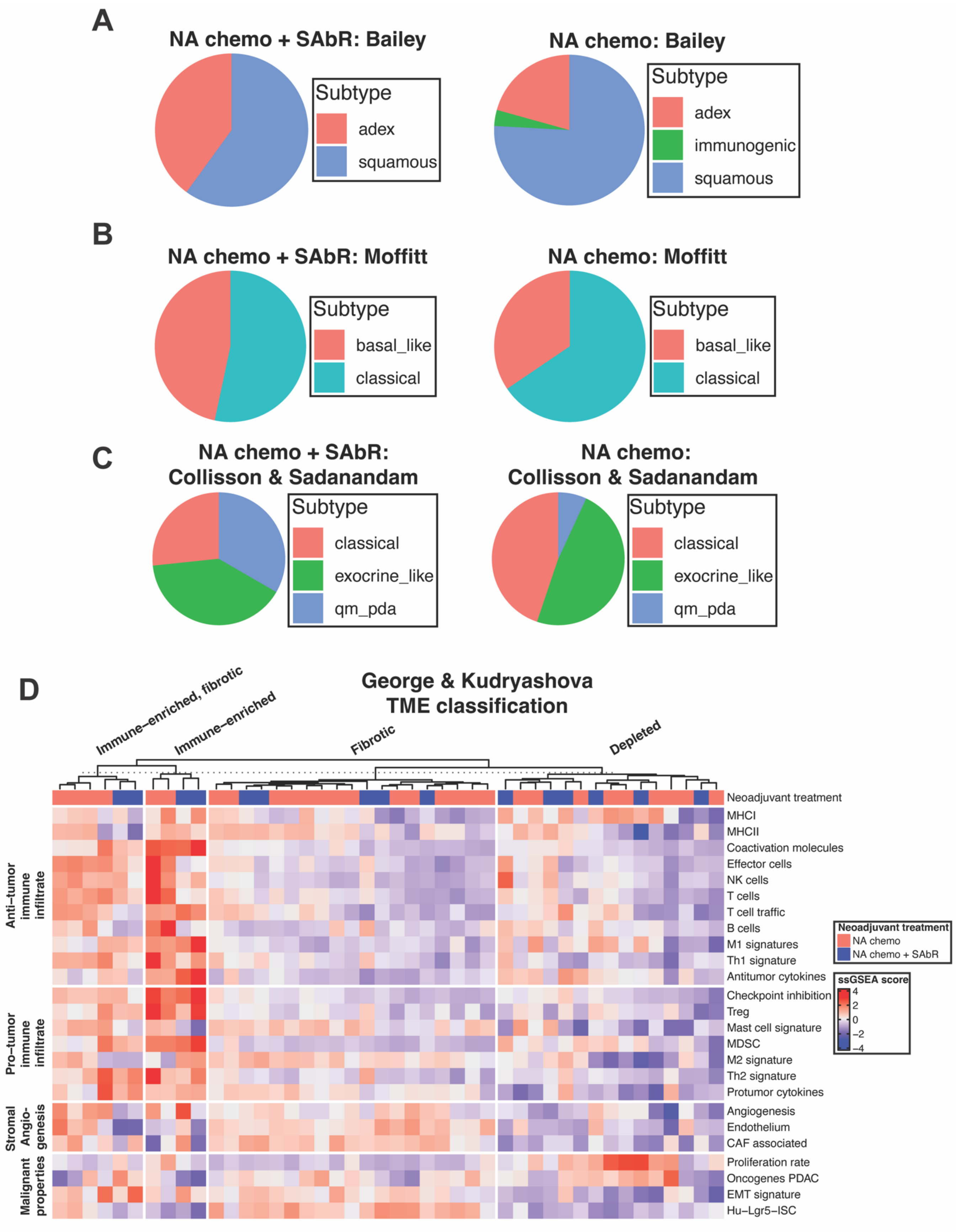


***Supplemental Figure 6: Molecular subtyping of pancreatic tumor samples based on public gene signatures.***

**(A)** Molecular subtyping based on public gene signatures derived from Bailey 2016. Top 50 differentially upregulated genes for each subtype vs all others with the lowest adjusted *P* values were used to define each subtype. ssGSEA score was calculated for each sample, and the sample was then assigned to the highest scoring subtype. **(B)** Subtyping based on Moffit 2015 signature. All genes that scored the highest for each of the two subtypes were used as a signature for their respective subtype. ssGSEA scores were calculated from these gene sets and samples were assigned to whichever score was higher. **(C)** Subtyping based on the 62 assigner genes from Collisson & Sadanandam 2011. ssGSEA was calculated and samples were assigned to highest subtype score. **(D)** George & Kudryashova 2024 TME subtyping. ssGSEA scores were calculated for the pre-defined 25 functional gene sets, scaled, then z-score normalized before hierarchical clustering into 4 groups. Fisher's Exact test comparison of proportions revealed no significant differences between subtypes assigned to NA SAbR + Chemo samples compared to those treated with NA Chemo only (Bailey: $P = 0.4344$, Moffit: $P = 0.5207$, Collisson & Sadanandam: $P = 0.07802$, George & Kudryashova: $P = 0.7133$).

**Univariable distant recurrence free survival Cox models for select variables**

| Variable | N | Hazard ratio | HR (95% CI) | *P* |
|---|---|---|---|---|
| **Lymphovascular invasion** | | | | |
| Not identified | 60 | | Reference | |
| Present | 120 | | 0.91 (0.57 – 1.43) | .67 |
| **N stage: post–treatment (ypN)** | | | | |
| No nodal involvement | 83 | | Reference | |
| Nodal involvement | 97 | | 1.75 (1.11 – 2.75) | .02 |
| **Neoadjuvant treatment** | | | | |
| Neoadjuvant chemo | 133 | | Reference | |
| Neoadjuvant chemo + SAbR | 48 | | 0.75 (0.44 – 1.28) | .29 |
| **T stage: post–treatment (ypT)** | | | | |
| T1: <=2cm greatest dimension | 56 | | Reference | |
| T2: >2cm, <=4 cm greatest dimension | 79 | | 1.47 (0.83 – 2.63) | .19 |
| T3: >4cm greatest dimension | 38 | | 2.82 (1.53 – 5.21) | <.001 |
| **Treatment effect** | | | | |
| Complete/near complete response, score 0/1 | 20 | | Reference | |
| Partial response, score 2 | 121 | | 2.85 (1.03 – 7.89) | .04 |
| Poor or no response, score 3 | 20 | | 4.39 (1.37 – 14.11) | .01 |

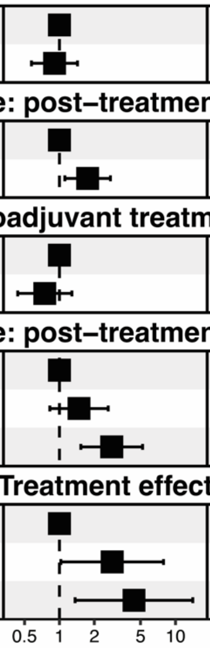


***Supplemental Figure 7: Univariable distant recurrence free survival Cox models for neoadjuvant treatment regimen, ypN, ypT, and treatment effect.*** Univariable cox models for distant metastasis progression free survival for neoadjuvant treatment regimen select pathological outcomes for all patients reported by pathologist based on CAP "Pancreas (Exocrine)" template. P-values from Wald test.

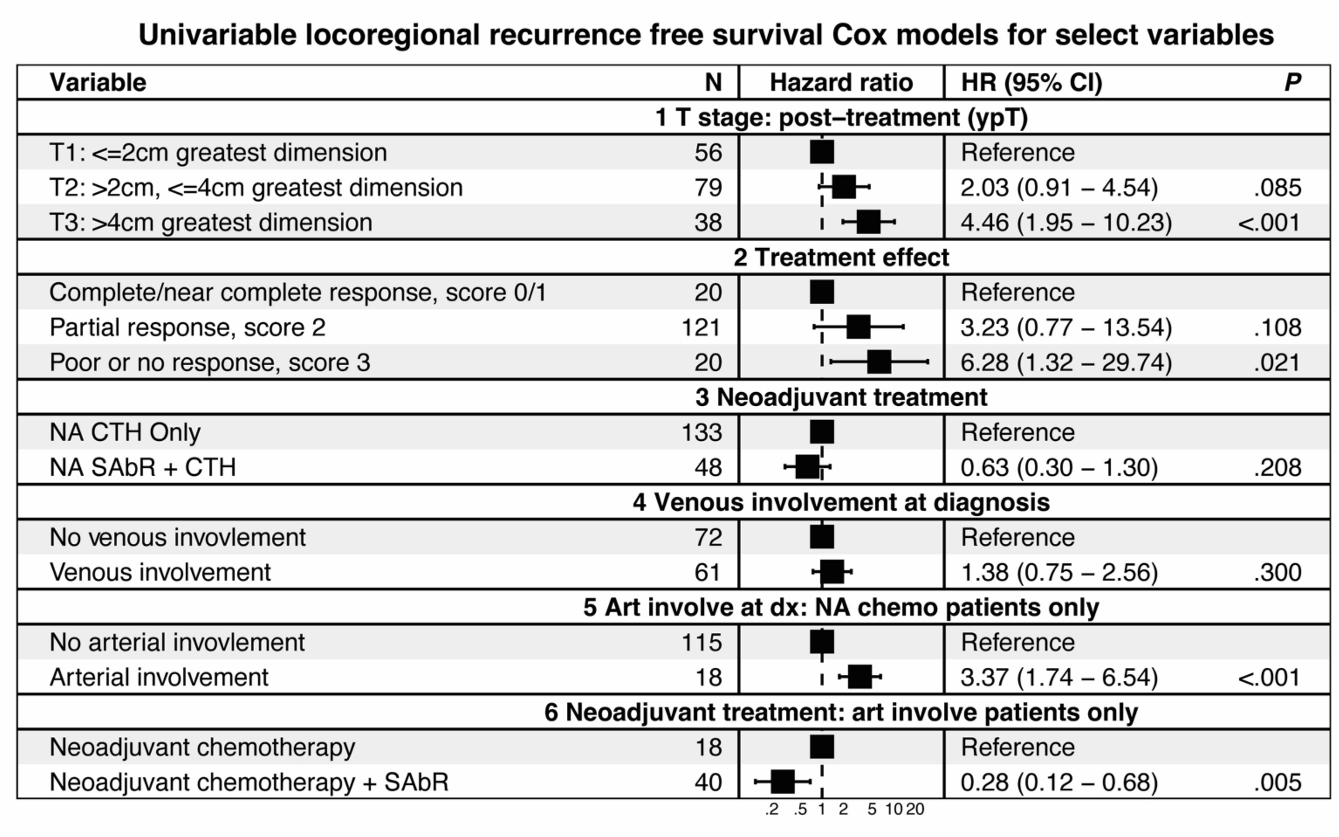

**Univariable locoregional recurrence free survival Cox models for select variables**

| Variable | N | Hazard ratio | HR (95% CI) | *P* |
|---|---|---|---|---|
| **1 T stage: post–treatment (ypT)** | | | | |
| T1: <=2cm greatest dimension | 56 | | Reference | |
| T2: >2cm, <=4cm greatest dimension | 79 | | 2.03 (0.91 – 4.54) | .085 |
| T3: >4cm greatest dimension | 38 | | 4.46 (1.95 – 10.23) | <.001 |
| **2 Treatment effect** | | | | |
| Complete/near complete response, score 0/1 | 20 | | Reference | |
| Partial response, score 2 | 121 | | 3.23 (0.77 – 13.54) | .108 |
| Poor or no response, score 3 | 20 | | 6.28 (1.32 – 29.74) | .021 |
| **3 Neoadjuvant treatment** | | | | |
| NA CTH Only | 133 | | Reference | |
| NA SAbR + CTH | 48 | | 0.63 (0.30 – 1.30) | .208 |
| **4 Venous involvement at diagnosis** | | | | |
| No venous invovlement | 72 | | Reference | |
| Venous involvement | 61 | | 1.38 (0.75 – 2.56) | .300 |
| **5 Art involve at dx: NA chemo patients only** | | | | |
| No arterial invovlement | 115 | | Reference | |
| Arterial involvement | 18 | | 3.37 (1.74 – 6.54) | <.001 |
| **6 Neoadjuvant treatment: art involve patients only** | | | | |
| Neoadjuvant chemotherapy | 18 | | Reference | |
| Neoadjuvant chemotherapy + SAbR | 40 | | 0.28 (0.12 – 0.68) | .005 |



***Supplemental Figure 8: Univariable locoregional recurrence free survival Cox models for select features.***

Univariable cox models with hazard ratios (HR), 95% confidence intervals (CI), and Wald-test *P* values for ypT, treatment effect, neoadjuvant treatment regimen, venous involvement of the tumor at diagnosis, arterial involvement of the tumor at diagnosis for NA chemo patients only, and neoadjuvant treatment for arterial-involved patients only.

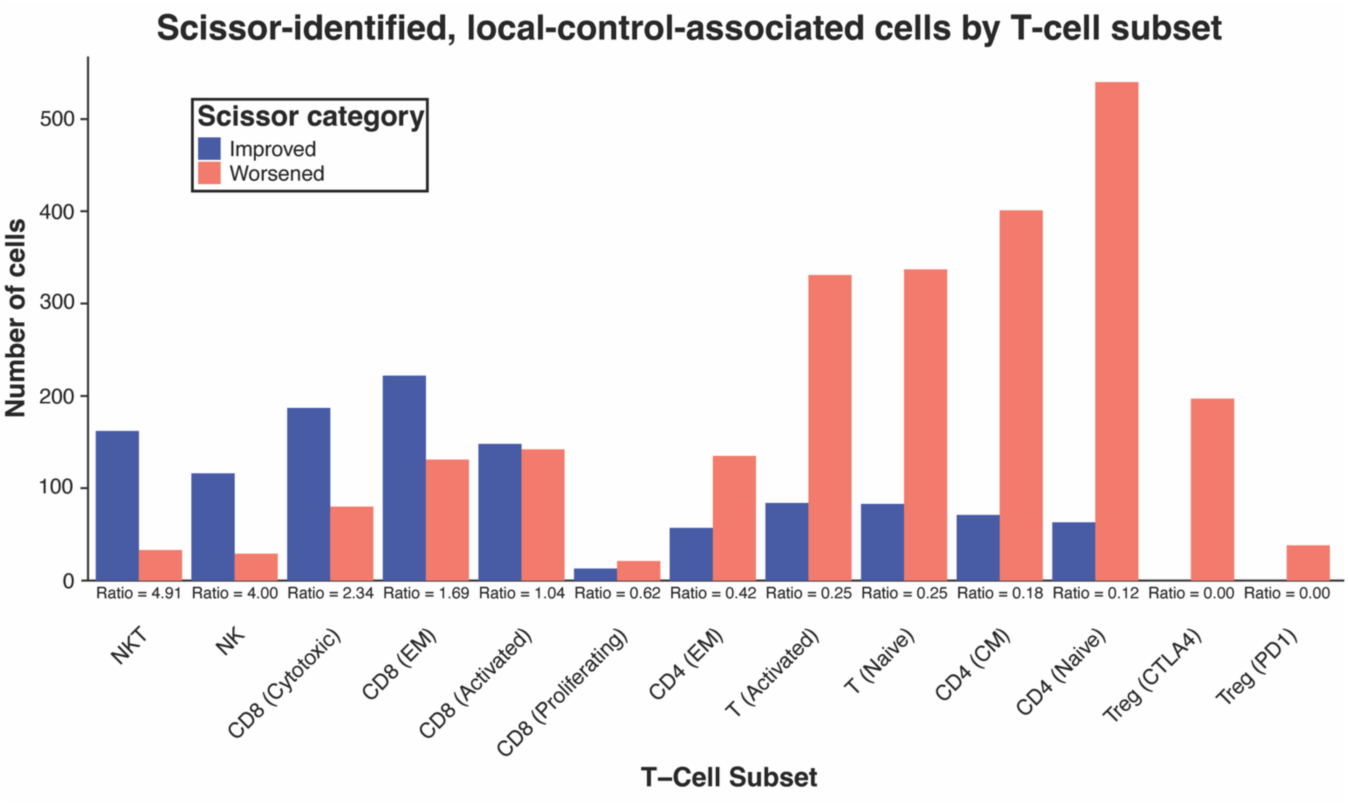


***Supplemental Figure 9: Detailed analysis of Scissor-identified in cells T-cell-subset associated with local control.*** Number of Scissor-identified cells in associated with local control. Cells are from high-resolution T cell subsets generated from public single-cell RNA sequencing PDAC datasets as in Figures 5C and 6B. Ratios of improved vs. worsened local control scissor-identified cells are provided, and subsets are ordered in descending order of improved to worsened ratio.

**Supplemental Table 1. Pre-surgical characteristics by neoadjuvant treatment**

| Characteristic | **NA chemo**, N = 133[1] | **NA chemo + SAbR**, N = 48[1] | ***P***[2] |
|---|---|---|---|
| **Age at diagnosis**[3] | | | .21 |
| Mean (SD) | 66 (9) | 64 (11) | |
| **Sex** | | | .87 |
| Male | 74 / 133 (56%) | 28 / 48 (58%) | |
| Female | 59 / 133 (44%) | 20 / 48 (42%) | |
| **NCCN resectability status at diagnosis** | | | **<.001** |
| Resectable | 85 / 133 (64%) | 2 / 48 (4%) | |
| Borderline resectable | 44 / 133 (33%) | 28 / 48 (58%) | |
| Locally advanced | 4 / 133 (3%) | 18 / 48 (38%) | |
| **Arterial involvement at diagnosis** | | | **<.001** |
| Involvement | 18 / 133 (14%) | 40 / 48 (83%) | |
| No involvement | 115 / 133 (86%) | 8 / 48 (17%) | |
| **T stage: pre-treatment (cT)** | | | **<.001** |
| 1 | 20 / 131 (15%) | 1 / 48 (2.1%) | |
| 2 | 75 / 131 (57%) | 6 / 48 (13%) | |
| 3 | 18 / 131 (14%) | 1 / 48 (2.1%) | |
| 4 | 18 / 131 (14%) | 40 / 48 (83%) | |
| **Tumor largest dimension at diagnosis (cm)**[4] | | | .87 |
| Mean (SD) | 3.20 (2.08) | 3.24 (1.10) | |
| **Tumor largest dimension post-treatment (cm)**[5] | | | .76 |
| Mean (SD) | 2.47 (1.25) | 2.54 (0.96) | |
| **Neoadjuvant chemotherapy regimen** | | | .71 |
| FOLFIRINOX | 104 / 132 (79%) | 41 / 48 (85%) | |
| Gemcitabine + Abraxane | 24 / 132 (18%) | 6 / 48 (13%) | |
| Other chemotherapy | 4 / 132 (3%) | 1 / 48 (2%) | |
| **Time on chemotherapy (months)**[6] | | | **.002** |
| Mean (SD) | 2.96 (1.00) | 4.47 (3.09) | |
| **Time from diagnosis to surgery (months)**[3] | | | **<.001** |
| Mean (SD) | 5.27 (2.14) | 9.25 (4.66) | |
| **Neoadjuvant radiation total dose (Gy)** | | | |
| Median (Range) | NA | 40 (24, 55) | |
| **Neoadjuvant radiation fractions**[7] | | | |
| Median (Range) | NA | 5 (3, 5) | |
| **Pre-treatment CA19-9 (U/mL)**[8] | | | .26 |
| Median (SD) | 288 (11715) | 359 (10214) | |
| **Post-chemo CA19-9 (U/mL)**[8] | | | .34 |
| Median (SD) | 53 (3663) | 21 (336) | |
| **Pre-surgery CA19-9 (U/mL)**[9] | | | .30 |
| Median (SD) | 54 (3663) | 23 (288) | |
| **Post-surgery CA19-9 (U/mL)**[10] | | | .63 |
| Median (SD) | 16 (423) | 31 (442) | |

[1] n / N (%)

[2] Welch Two Sample t-test; Fisher's exact test

[3] Representative of 133 NA chemo and 48 NA chemo + SAbR patients

[4] Representative of 131 NA chemo and 47 NA chemo + SAbR patients

[5] Representative of 105 NA chemo and 34 NA chemo + SAbR patients

[6] Representative of 124 NA chemo and 48 NA chemo + SAbR patients

[7] Representative of 0 NA chemo and 48 NA chemo + SAbR patients

[8] Representative of 77 NA chemo and 28 NA chemo + SAbR patients

[9] Representative of 67 NA chemo and 40 NA chemo + SAbR patients

[10] Representative of 65 NA chemo and 25 NA chemo + SAbR patients

**Supplemental Table 2. Pre-surgical characteristics: all NA chemo + SAbR patients vs RNA-sequenced subset**

| Characteristic | **All NA chemo + SAbR**, N = 48[1] | **RNA-sequenced subset**, N = 14[1] | ***P***[2] |
|---|---|---|---|
| **Age at diagnosis**[3] | | | .33 |
| Mean (SD) | 64 (11) | 67 (10) | |
| **Sex** | | | .22 |
| Male | 28 / 48 (58%) | 11 / 14 (79%) | |
| Female | 20 / 48 (42%) | 3 / 14 (21%) | |
| **NCCN resectability status at diagnosis** | | | .89 |
| Resectable | 2 / 48 (4%) | 1 / 14 (7%) | |
| Borderline resectable | 28 / 48 (58%) | 8 / 14 (57%) | |
| Locally advanced | 18 / 48 (38%) | 5 / 14 (36%) | |
| **Arterial involvement at diagnosis** | | | .70 |
| Involvement | 40 / 48 (83%) | 11 / 14 (79%) | |
| No involvement | 8 / 48 (17%) | 3 / 14 (21%) | |
| **T stage: pre-treatment (cT)** | | | .66 |
| 1 | 1 / 48 (2.1%) | 0 / 14 (0%) | |
| 2 | 6 / 48 (13%) | 3 / 14 (21%) | |
| 3 | 1 / 48 (2.1%) | 0 / 14 (0%) | |
| 4 | 40 / 48 (83%) | 11 / 14 (79%) | |
| **Tumor largest dimension at diagnosis (cm)**[4] | | | .38 |
| Mean (SD) | 3.24 (1.10) | 3.00 (0.80) | |
| **Tumor largest dimension post-treatment (cm)**[5] | | | .66 |
| Mean (SD) | 2.54 (0.96) | 2.39 (0.89) | |
| **Neoadjuvant chemotherapy regimen** | | | .63 |
| FOLFIRINOX | 41 / 48 (85%) | 11 / 14 (79%) | |
| Gemcitabine + Abraxane | 6 / 48 (13%) | 2 / 14 (14%) | |
| Other chemotherapy | 1 / 48 (2%) | 1 / 14 (7%) | |
| **Time on chemotherapy (months)**[3] | | | .23 |
| Mean (SD) | 4.47 (3.09) | 3.71 (1.62) | |
| **Time from diagnosis to surgery (months)**[3] | | | .48 |
| Mean (SD) | 9.3 (4.7) | 8.5 (3.1) | |
| **Neoadvjuvant radiation total dose (Gy)**[3] | | | .63 |
| Median (Range) | 40 (24, 55) | 40 (35, 50) | |
| **Neoadjuvant radiation fractions**[3] | | | .32 |
| Median (Range) | 5 (3, 5) | 5 (5, 5) | |
| **Pre-treatment CA19-9 (U/mL)**[6] | | | .82 |
| Median (SD) | 359 (10,214) | 2,164 (12,622) | |
| **Post-chemo CA19-9 (U/mL)**[6] | | | .177 |
| Median (SD) | 21 (336) | 26 (96) | |
| **Pre-surgery CA19-9 (U/mL)**[7] | | | .33 |
| Median (SD) | 23 (288) | 21 (108) | |
| **Post-surgery CA19-9 (U/mL)**[8] | | | .68 |
| Median (SD) | 31 (442) | 34 (689) | |

[1] n / N (%)
[2] Welch Two Sample t-test; Fisher's exact test
[3] Representative of 48 of all NA chemo + SAbR and 14 RNA-sequenced NA chemo + SAbR patients
[4] Representative of 47 of all NA chemo + SAbR and 13 RNA-sequenced NA chemo + SAbR patients
[5] Representative of 34 of all NA chemo + SAbR and 10 RNA-sequenced NA chemo + SAbR patients
[6] Representative of 28 of all NA chemo + SAbR and 8 RNA-sequenced NA chemo + SAbR patients
[7] Representative of 40 of all NA chemo + SAbR and 11 RNA-sequenced NA chemo + SAbR patients
[8] Representative of 25 of all NA chemo + SAbR and 6 RNA-sequenced NA chemo + SAbR patients

### Supplemental Table 3. Pre-surgical characteristics: all NA chemo patients vs RNA-sequenced subset

| Characteristic | **All NA chemo**, N = 133[1] | **RNA-sequenced subset**, N = 29[1] | ***P***[2] |
|---|---|---|---|
| **Age at diagnosis**[3] | | | .130 |
| Mean (SD) | 66 (9) | 64 (9) | |
| **Sex** | | | .84 |
| Male | 74 / 133 (56%) | 15 / 29 (52%) | |
| Female | 59 / 133 (44%) | 14 / 29 (48%) | |
| **NCCN resectability status at diagnosis** | | | .47 |
| Resectable | 85 / 133 (64%) | 22 / 29 (76%) | |
| Borderline resectable | 44 / 133 (33%) | 7 / 29 (24%) | |
| Locally advanced | 4 / 133 (3%) | 0 / 29 (0%) | |
| **Arterial involvement at diagnosis** | | | >.9 |
| Involvement | 18 / 133 (14%) | 4 / 29 (14%) | |
| No involvement | 115 / 133 (86%) | 25 / 29 (86%) | |
| **T stage: pre-treatment (cT)** | | | .72 |
| 1 | 20 / 131 (15%) | 2 / 29 (6.9%) | |
| 2 | 75 / 131 (57%) | 18 / 29 (62%) | |
| 3 | 18 / 131 (14%) | 5 / 29 (17%) | |
| 4 | 18 / 131 (14%) | 4 / 29 (14%) | |
| **Tumor largest dimension at diagnosis (cm)**[4] | | | .77 |
| Mean (SD) | 3.20 (2.08) | 3.29 (1.16) | |
| **Tumor largest dimension post-treatment (cm)**[5] | | | >.9 |
| Mean (SD) | 2.47 (1.25) | 2.46 (1.01) | |
| **Neoadjuvant chemotherapy regimen** | | | .32 |
| FOLFIRINOX | 104 / 132 (79%) | 26 / 28 (93%) | |
| Gemcitabine + Abraxane | 24 / 132 (18%) | 2 / 28 (7%) | |
| Other chemotherapy | 4 / 132 (3%) | 0 / 28 (0%) | |
| **Time on chemotherapy (months)**[6] | | | .113 |
| Mean (SD) | 2.96 (1.00) | 2.66 (0.84) | |
| **Time from diagnosis to surgery (months)**[3] | | | .89 |
| Mean (SD) | 5.27 (2.14) | 5.36 (2.84) | |
| **Pre-treatment CA19-9 (U/mL)**[7] | | | .49 |
| Median (SD) | 288 (11,715) | 192 (24,739) | |
| **Post-chemo CA19-9 (U/mL)**[7] | | | .51 |
| Median (SD) | 53 (3,663) | 70 (7,234) | |
| **Pre-surgery CA19-9 (U/mL)**[8] | | | .50 |
| Median (SD) | 54 (3,663) | 70 (7,234) | |
| **Post-surgery CA19-9 (U/mL)**[9] | | | >.9 |
| Median (SD) | 16 (423) | 16 (436) | |

[1] n / N (%)
[2] Welch Two Sample t-test; Fisher's exact test
[3] Representative of 133 of all NA chemo + SAbR and 29 RNA-sequenced NA chemo + SAbR patients
[4] Representative of 131 of all NA chemo + SAbR and 29 RNA-sequenced NA chemo + SAbR patients
[5] Representative of 105 of all NA chemo + SAbR and 25 RNA-sequenced NA chemo + SAbR patients
[6] Representative of 124 of all NA chemo + SAbR and 28 RNA-sequenced NA chemo + SAbR patients
[7] Representative of 77 of all NA chemo + SAbR and 17 RNA-sequenced NA chemo + SAbR patients
[8] Representative of 67 of all NA chemo + SAbR and 17 RNA-sequenced NA chemo + SAbR patients
[9] Representative of 65 of all NA chemo + SAbR and 15 RNA-sequenced NA chemo + SAbR patients

### Supplemental Table 4. Baseline characteristics of matched patients by neoadjuvant treatment

| Characteristic | **NA chemo**, N = 25[1] | **NA chemo + SAbR**, N = 25[1] | ***P***[2] |
|---|---|---|---|
| **Age at diagnosis**[3] | | | .83 |
| Mean (SD) | 66 (9) | 66 (10) | |
| **Sex** | | | .77 |
| Male | 17 / 25 (68%) | 15 / 25 (60%) | |
| Female | 8 / 25 (32%) | 10 / 25 (40%) | |
| **NCCN resectability status at diagnosis** | | | >.9 |
| Resectable | 2 / 25 (8%) | 2 / 25 (8%) | |
| Borderline resectable | 20 / 25 (80%) | 20 / 25 (80%) | |
| Locally advanced | 3 / 25 (12%) | 3 / 25 (12%) | |
| **Arterial involvement at diagnosis** | | | >.9 |
| Involvement | 16 / 25 (64%) | 17 / 25 (68%) | |
| No involvement | 9 / 25 (36%) | 8 / 25 (32%) | |
| **Tumor largest dimension at diagnosis (cm)**[3] | | | >.9 |
| Mean (SD) | 3.10 (1.63) | 3.08 (0.91) | |
| **T stage: pre-treatment (cT)** | | | >.9 |
| 1 | 1 / 25 (4.0%) | 1 / 25 (4.0%) | |
| 2 | 7 / 25 (28%) | 6 / 25 (24%) | |
| 3 | 1 / 25 (4.0%) | 1 / 25 (4.0%) | |
| 4 | 16 / 25 (64%) | 17 / 25 (68%) | |
| **Neoadjuvant chemotherapy regimen** | | | .31 |
| FOLFIRINOX | 19 / 25 (76%) | 21 / 25 (84%) | |
| Gemcitabine + Abraxane | 3 / 25 (12%) | 4 / 25 (16%) | |
| Other chemotherapy | 3 / 25 (12%) | 0 / 25 (0%) | |
| **Time on chemotherapy (months)**[3] | | | .74 |
| Mean (SD) | 3.22 (1.42) | 3.37 (1.73) | |

[1] n / N (%)

[2] Welch Two Sample t-test; Fisher's exact test

[3] Representative of 25 NA chemo and 25 NA chemo + SAbR patients

### Supplemental Table 5. Post-surgical pathological characteristics by neoadjuvant treatment

| Characteristic | **NA chemo**, N = 133[1] | **NA chemo + SAbR**, N = 48[1] | ***P***[2] |
|---|---|---|---|
| **Tumor site** | | | .162 |
| Head/neck | 106 / 133 (80%) | 33 / 48 (69%) | |
| Tail | 27 / 133 (20%) | 15 / 48 (31%) | |
| **Margin status** | | | .40 |
| Involved | 28 / 131 (21%) | 7 / 48 (15%) | |
| Uninvolved | 103 / 131 (79%) | 41 / 48 (85%) | |
| **Lymphovascular invasion** | | | .107 |
| Present | 93 / 132 (70%) | 27 / 48 (56%) | |
| Not identified | 39 / 132 (30%) | 21 / 48 (44%) | |
| **Perineural invasion** | | | **.019** |
| Present | 105 / 131 (80%) | 30 / 48 (63%) | |
| Not identified | 26 / 131 (20%) | 18 / 48 (38%) | |
| **Histological grade** | | | .058 |
| G1: Well differentiated | 6 / 121 (5%) | 0 / 43 (0%) | |
| G2: Moderately differentiated | 80 / 121 (66%) | 23 / 43 (53%) | |
| G3: Poorly differentiated | 35 / 121 (29%) | 20 / 43 (47%) | |
| **Treatment effect** | | | **<.001** |
| Complete/near complete response, score 0/1 | 8 / 113 (7%) | 12 / 48 (25%) | |
| Partial response, score 2 | 85 / 113 (75%) | 36 / 48 (75%) | |
| Poor or no response, score 3 | 20 / 113 (18%) | 0 / 48 (0%) | |
| **T stage: post-treatment (ypT)** | | | **.002** |
| T0: No evidence of primary tumor | 1 / 130 (1%) | 1 / 48 (2%) | |
| T1: Tumor ≤2cm in greatest dimension | 35 / 130 (27%) | 21 / 48 (44%) | |
| T2: Tumor >2cm and ≤4 cm in greatest dimension | 61 / 130 (47%) | 18 / 48 (38%) | |
| T3: Tumor >4cm In greatest dimension | 33 / 130 (25%) | 5 / 48 (10%) | |
| T4: Tumor Involves the celiac axis, superior mesenteric artery, and/or common hepatic artery | 0 / 130 (0%) | 3 / 48 (6%) | |
| **N Stage: Post-Treatment (ypN)** | | | **.015** |
| N0: No regional lymph node metastasis | 54 / 132 (41%) | 29 / 48 (60%) | |
| N1: Metastasis in one to three regional lymph nodes | 54 / 132 (41%) | 17 / 48 (35%) | |
| N2: Metastasis in four or more regional lymph nodes | 24 / 132 (18%) | 2 / 48 (4%) | |

[1] n / N (%)

[2] Fisher's exact test

**Supplemental Table 6. Post-surgical pathological characteristics: NA chemo + SAbR patients vs RNA sequenced subset**

| Characteristic | All NA chemo + SAbR Patients, N = 48[1] | RNA-seq NA chemo + SAbR Patients, N = 16[1] | *P*[2] |
|---|---|---|---|
| **Tumor site** | | | .23 |
| Head/neck | 33 / 48 (69%) | 8 / 16 (50%) | |
| Tail | 15 / 48 (31%) | 8 / 16 (50%) | |
| **Margin status** | | | >.9 |
| Involved | 7 / 48 (15%) | 2 / 16 (13%) | |
| Uninvolved | 41 / 48 (85%) | 14 / 16 (88%) | |
| **Lymphovascular invasion** | | | .24 |
| Present | 27 / 48 (56%) | 12 / 16 (75%) | |
| Not identified | 21 / 48 (44%) | 4 / 16 (25%) | |
| **Perineural invasion** | | | .77 |
| Present | 30 / 48 (63%) | 9 / 16 (56%) | |
| Not identified | 18 / 48 (38%) | 7 / 16 (44%) | |
| **Histological grade** | | | .77 |
| G1: Well differentiated | 0 / 43 (0%) | 0 / 15 (0%) | |
| G2: Moderately differentiated | 23 / 43 (53%) | 9 / 15 (60%) | |
| G3: Poorly differentiated | 20 / 43 (47%) | 6 / 15 (40%) | |
| **Treatment effect** | | | .74 |
| Complete/near complete response, score 0/1 | 12 / 48 (25%) | 3 / 16 (19%) | |
| Partial response, score 2 | 36 / 48 (75%) | 13 / 16 (81%) | |
| Poor or no response, score 3 | 0 / 48 (0%) | 0 / 16 (0%) | |
| **T stage: post-treatment (ypT)** | | | .59 |
| T0: No evidence of primary tumor | 1 / 48 (2%) | 0 / 16 (0%) | |
| T1: Tumor ≤2cm in greatest dimension | 21 / 48 (44%) | 10 / 16 (63%) | |
| T2: Tumor >2cm and ≤4 cm in greatest dimension | 18 / 48 (38%) | 3 / 16 (19%) | |
| T3: Tumor >4cm In greatest dimension | 5 / 48 (10%) | 2 / 16 (13%) | |
| T4: Tumor Involves the celiac axis, superior mesenteric artery, and/or common hepatic artery | 3 / 48 (6%) | 1 / 16 (6%) | |
| **N stage: post-treatment (ypN)** | | | .87 |
| N0: No regional lymph node metastasis | 29 / 48 (60%) | 9 / 16 (56%) | |
| N1: Metastasis in one to three regional lymph nodes | 17 / 48 (35%) | 7 / 16 (44%) | |
| N2: Metastasis in four or more regional lymph nodes | 2 / 48 (4%) | 0 / 16 (0%) | |

[1] n / N (%)

[2] Fisher's exact test

**Supplemental Table 7. Post-surgical pathological characteristics: NA chemo patients vs RNA sequenced subset**

| Characteristic | All NA chemo Patients, N = 133[1] | RNA-seq NA chemo Patients, N = 34[1] | P[2] |
|---|---|---|---|
| **Tumor site** | | | >.9 |
| Head/neck | 106 / 133 (80%) | 27 / 34 (79%) | |
| Tail | 27 / 133 (20%) | 7 / 34 (21%) | |
| **Margin status** | | | .81 |
| Involved | 28 / 131 (21%) | 6 / 34 (18%) | |
| Uninvolved | 103 / 131 (79%) | 28 / 34 (82%) | |
| **Lymphovascular invasion** | | | .39 |
| Present | 93 / 132 (70%) | 27 / 34 (79%) | |
| Not identified | 39 / 132 (30%) | 7 / 34 (21%) | |
| **Perineural invasion** | | | .64 |
| Present | 105 / 131 (80%) | 26 / 34 (76%) | |
| Not identified | 26 / 131 (20%) | 8 / 34 (24%) | |
| **Histological grade** | | | .37 |
| G1: Well differentiated | 6 / 121 (5%) | 2 / 30 (7%) | |
| G2: Moderately differentiated | 80 / 121 (66%) | 16 / 30 (53%) | |
| G3: Poorly differentiated | 35 / 121 (29%) | 12 / 30 (40%) | |
| **Treatment effect** | | | .73 |
| Complete/near complete response, score 0/1 | 8 / 113 (7%) | 3 / 32 (9%) | |
| Partial response, score 2 | 85 / 113 (75%) | 25 / 32 (78%) | |
| Poor or no response, score 3 | 20 / 113 (18%) | 4 / 32 (13%) | |
| **T stage: post-treatment (ypT)** | | | .59 |
| T0: No evidence of primary tumor | 1 / 130 (1%) | 0 / 34 (0%) | |
| T1: Tumor ≤2cm in greatest dimension | 35 / 130 (27%) | 8 / 34 (24%) | |
| T2: Tumor >2cm and ≤4 cm in greatest dimension | 61 / 130 (47%) | 20 / 34 (59%) | |
| T3: Tumor >4cm In greatest dimension | 33 / 130 (25%) | 6 / 34 (18%) | |
| T4: Tumor Involves the celiac axis, superior mesenteric artery, and/or common hepatic artery | 0 / 130 (0%) | 0 / 34 (0%) | |
| **N stage: post-treatment (ypN)** | | | .37 |
| N0: No regional lymph node metastasis | 54 / 132 (41%) | 12 / 34 (35%) | |
| N1: Metastasis in one to three regional lymph nodes | 54 / 132 (41%) | 12 / 34 (35%) | |
| N2: Metastasis in four or more regional lymph nodes | 24 / 132 (18%) | 10 / 34 (29%) | |

[1] n / N (%)

[2] Fisher's exact test

### Supplemental Table 8. Pre-surgical characteristics: NA chemo patients by arterial involvement

| Characteristic | **Arterial involvement**, N = 18[1] | **No involvement**, N = 115[1] | $P^2$ |
|---|---|---|---|
| **Age at diagnosis**[3] | | | >.9 |
| Mean (SD) | 66 (8) | 66 (9) | |
| **Sex** | | | >.9 |
| Male | 10 / 18 (56%) | 64 / 115 (56%) | |
| Female | 8 / 18 (44%) | 51 / 115 (44%) | |
| **NCCN resectability status at diagnosis** | | | **<.001** |
| Resectable | 1 / 18 (6%) | 84 / 115 (73%) | |
| Borderline resectable | 14 / 18 (78%) | 30 / 115 (26%) | |
| Locally advanced | 3 / 18 (17%) | 1 / 115 (1%) | |
| **T stage: pre-treatment (cT)** | | | **<.001** |
| 1 | 0 / 18 (0%) | 20 / 113 (18%) | |
| 2 | 0 / 18 (0%) | 75 / 113 (66%) | |
| 3 | 0 / 18 (0%) | 18 / 113 (16%) | |
| 4 | 18 / 18 (100%) | 0 / 113 (0%) | |
| **Tumor largest dimension at diagnosis (cm)**[4] | | | .87 |
| Mean (SD) | 3.27 (1.80) | 3.19 (2.13) | |
| **Tumor largest dimension post-treatment (cm)**[5] | | | .129 |
| Mean (SD) | 3.04 (1.52) | 2.38 (1.18) | |
| **Neoadjuvant chemotherapy regimen** | | | .54 |
| FOLFIRINOX | 15 / 18 (83%) | 89 / 114 (78%) | |
| Gemcitabine + Abraxane | 2 / 18 (11%) | 22 / 114 (19%) | |
| Other chemotherapy | 1 / 18 (6%) | 3 / 114 (3%) | |
| **Time on chemotherapy (months)**[6] | | | .85 |
| Mean (SD) | 2.90 (1.22) | 2.96 (0.97) | |
| **Time from diagnosis to surgery (months)**[3] | | | .63 |
| Mean (SD) | 5.07 (1.81) | 5.31 (2.20) | |
| **Pre-treatment CA19-9 (U/mL)**[7] | | | .89 |
| Median (SD) | 366 (3,608) | 286 (12,330) | |
| **Post-chemo CA19-9 (U/mL)**[7] | | | .23 |
| Median (SD) | 28 (42) | 57 (3,901) | |
| **Pre-surgery CA19-9 (U/mL)**[8] | | | .23 |
| Median (SD) | 32 (41) | 57 (3,901) | |
| **Post-surgery CA19-9 (U/mL)**[9] | | | .062 |
| Median (SD) | 15 (50) | 17 (454) | |

[1] n / N (%)
[2] Welch Two Sample t-test; Fisher's exact test
[3] Representative of 18 NA chemo patients with arterial invovlement and 115 without
[4] Representative of 18 NA chemo patients with arterial invovlement and 113 without
[5] Representative of 15 NA chemo patients with arterial invovlement and 90 without
[6] Representative of 16 NA chemo patients with arterial invovlement and 108 without
[7] Representative of 8 NA chemo patients with arterial invovlement and 69 without
[8] Representative of 8 NA chemo patients with arterial invovlement and 59 without
[9] Representative of 9 NA chemo patients with arterial invovlement and 56 without

**Supplemental Table 9. Pre-surgical characteristics: arterial involved patients by neoadj treatment**

| Characteristic | **NA chemo**, N = 18[1] | **NA chemo + SAbR**, N = 40[1] | ***P***[2] |
|---|---|---|---|
| **Age at diagnosis**[3] | | | .40 |
| Mean (SD) | 66 (8) | 64 (10) | |
| **Sex** | | | >.9 |
| Male | 10 / 18 (56%) | 22 / 40 (55%) | |
| Female | 8 / 18 (44%) | 18 / 40 (45%) | |
| **NCCN resectability status at diagnosis** | | | **.034** |
| Resectable | 1 / 18 (6%) | 0 / 40 (0%) | |
| Borderline resectable | 14 / 18 (78%) | 22 / 40 (55%) | |
| Locally advanced | 3 / 18 (17%) | 18 / 40 (45%) | |
| **Tumor largest dimension at diagnosis (cm)**[4] | | | >.9 |
| Mean (SD) | 3.27 (1.80) | 3.28 (1.15) | |
| **Tumor largest dimension post-treatment (cm)**[5] | | | .33 |
| Mean (SD) | 3.04 (1.52) | 2.60 (1.02) | |
| **Neoadjuvant chemotherapy regimen** | | | .84 |
| FOLFIRINOX | 15 / 18 (83%) | 35 / 40 (88%) | |
| Gemcitabine + Abraxane | 2 / 18 (11%) | 4 / 40 (10%) | |
| Other chemotherapy | 1 / 18 (6%) | 1 / 40 (3%) | |
| **Time on chemotherapy (months)**[6] | | | **.007** |
| Mean (SD) | 2.90 (1.22) | 4.53 (3.15) | |
| **Time from diagnosis to surgery (months)**[3] | | | **<.001** |
| Mean (SD) | 5.07 (1.81) | 8.92 (4.55) | |
| **Neoadjuvant radiation total dose (Gy)** | | | |
| Median (Range) | NA | 40 (32, 55) | |
| **Neoadjuvant radiation fractions**[7] | | | |
| Median (Range) | NA | 5 (5, 5) | |
| **Pre-treatment CA19-9 (U/mL)**[8] | | | .35 |
| Median (SD) | 366 (3,608) | 197 (10,633) | |
| **Post-chemo CA19-9 (U/mL)**[8] | | | .109 |
| Median (SD) | 28 (42) | 19 (321) | |
| **Pre-surgery CA19-9 (U/mL)**[9] | | | .166 |
| Median (SD) | 32 (41) | 30 (153) | |
| **Post-surgery CA19-9 (U/mL)**[10] | | | .34 |
| Median (SD) | 15 (50) | 19 (192) | |

[1] n / N (%)
[2] Welch Two Sample t-test; Fisher's exact test
[3] Representative of 18 NA chemo and 40 NA chemo + SAbR patients with arterial involvement
[4] Representative of 18 NA chemo and 39 NA chemo + SAbR patients with arterial involvement
[5] Representative of 15 NA chemo and 28 NA chemo + SAbR patients with arterial involvement
[6] Representative of 16 NA chemo and 40 NA chemo + SAbR patients with arterial involvement
[7] Representative of 0 NA chemo and 40 NA chemo + SAbR patients with arterial involvement
[8] Representative of 8 NA chemo and 23 NA chemo + SAbR patients with arterial involvement
[9] Representative of 8 NA chemo and 33 NA chemo + SAbR patients with arterial involvement
[10] Representative of 9 NA chemo and 19 NA chemo + SAbR patients with arterial involvement
